# Modelling of COVID-19 pandemic vis-à-vis some socio-economic factors

**DOI:** 10.1101/2021.09.30.21264356

**Authors:** Kayode Oshinubi, Mustapha Rachdi, Jacques Demongeot

## Abstract

The impacts of COVID-19 outbreak on socio-economic status of countries across the globe cannot be overemphasized as we examine the role it played in various countries. A lot of people were out of jobs, many households were careful of their spending and a greater social fracture of the population in fourteen different countries has emerged. We considered periods of infection spread during the first and second wave in Organization for Economic Co-operation and Development (OECD) countries and countries in Africa, that is developed and developing countries alongside their social-economic data. We established a mathematical and statistical relationship between Theil and Gini index, then we studied the relationship between the data from epidemiology and socio-economic determinants using several machine learning and deep learning methods. High correlations were observed between some of the socio-economic and epidemiologic parameters and we predicted three of the socio-economic variables in order to validate our results. These result shows a sharp difference between the first and second wave of the pandemic confirming the real dynamics of the spread of the outbreak in several countries and ways by which it was mitigated.

## 1 Introduction

The modelling of COVID-19 by scientists, epidemiologists and health experts has been considered since the beginning of the pandemic as it ravages the world. Socio-economic determinants of this pandemic are important as they project how severe a country is hit and how it is being controlled, this leading to consider these variables alongside daily reproduction rates during the contagiousness period of individuals infected by the pandemic.

Some researchers have worked on socio-economic analysis of COVID-19 pandemic, they are as follows : in [1], the authors examined the geoclimatic, demographic and socio-economic determinants of COVID-19 prevalence and have shown that the influence of these determinants varies by comparing the first and second wave of the pandemic. The socio-economic impact of the COVID-19 pandemic in United State of America (USA) was studied by [2], where the authors investigate the systematic risk posted by sector-level industries within the USA. [3] modeled daily confirmed cases of COVID-19 in different countries across the globe using regression models with predictions for upcoming scenarios. [7] worked on the socio-economic and environmental factors influencing the basic reproduction number of COVID-19 pandemic by fitting a logistic growth curve to report daily cases up to the first peak of the pandemic while [8] studied the impacts of social and economic factors on the transmission of COVID-19 disease with China as a case study using an empirical model, and the authors conclude that these determinants have rich implications for ongoing efforts in containing the pandemic. The work in this present article is an extension of [16] which was based on the analysis of the reproduction number of COVID-19 based on the current health expenditure as Gross Domestic Product Percentage (CHE/GDP) across several countries using some machine learning tools.

Our goal in this article is to establish a relationship between Theil and Gini index, analyze critically some of the socio-economic determinants of the pandemic, correlate them, predict three of the socio-economic variables and perform some regression analysis. We also cluster countries according to these parameters and with the help of the lasso method (least absolute shrinkage and selection operator) we were able to select the best variables for this modelling.

The paper is divided into seven sections, in section two we explain the methodology used in this research. Section three deals with the variables used, in section four we established a mathematical and statistical relationship between Theil and Gini index, section five is dedicated to the visualization of the results obtained, while we finally gave the discussion and conclusion in section six and seven respectively.

## 2 Methods

The use of machine learning and deep learning methods to analyze data has been helpful over the years to get a proper view on how a model behaves. In this research we used some supervised and unsupervised machine learning methods and we also tried to use one deep learning method to see the visualization.

The supervised machine learning methods we used are polynomial regression, linear regression, lasso regression and ridge regression. We also use some of these methods to make prediction by training the model and testing some percentage of the values. Lasso regression helped us to know the best variables to be used in the modelling. We used unsupervised learning to cluster variables across countries and the methods we used to validate our results are K-means clustering, Hierarchy clustering and principal component analysis.

We also performed correlation among parameters used in this modelling and an optimization method called ordinary least square (OLS) to model the socio-economic determinants of COVID-19. The deep learning method we used are neural network and multilayer perceptron (MLP) regressor which is a class of feedforward artificial neural network (ANN).

Multivariate least square method allows us to test much more complex relations between variables. It can be can be represented as follows:

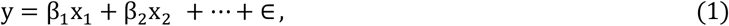

where β_1_, β_2_ … are coefficients or weights, ∈ is the residual noise, y is the dependent variable and x_1_, x_2_, … are the independent variables.

Ridge and lasso regression are simple methods to reduce the model complexity and prevent over-fitting which may result from linear regression. The cost function for ridge regression is given below:

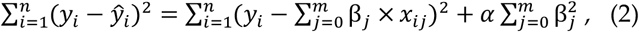

with for some 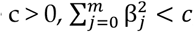, while α is the penalty term that regularizes the coefficients such that if the coefficients take large values, the optimization function is penalized. Ridge regression puts constraint on the coefficients β. We define the cost function for lasso regression as:

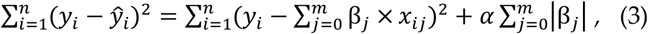

with for some 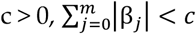.

## 3 Variables

### 3.1 Socio-economic Variables

Socio-economic variables are a strong determinant of the spread of the pandemic. We extracted some data from ([4-5] and [9-14]), while we also calculated some of the socio-economic variables.

The observed variables used in this research are immigration rate, average life expectancy (L.E), Tuberculosis incidence (TB), temperature, percentage of gross domestic product devoted to health expenditure (% GDP H. E) collated from [16], ten percentage lowest (L.I) and highest income (H.I), government response stringency index, sustainable development goal (SDG) index, human development index (HDI), environmental performance index (EPI), consumer confidence index (CCI), stringency index (S.I), Theil index (T.I) and Gini index (G.I). We collated the data based on the available countries and most recent years.

The calculated socio-economic variables are as follows:

– Social fracture (S.F) index which is the ratio between the ten percentage highest income and the ten percentage lowest income. In brief it is expressed by the formula below;

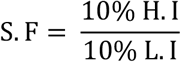
– Demographic Index (D.I) is the ratio between the percentage of GDP devoted to health expenditure and Social fracture index. It is expressed by equation below:

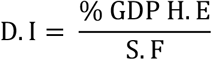

We give a precise values of all variables in Tables 3, 4 and 5 in the Appendix.

**Table 1.** Comparison of different regression models for the prediction.

| S/N | Country Name | Gini Index | Linear Regression | Lasso Regression | Ridge Regression | MLP Regressor |
| --- | --- | --- | --- | --- | --- | --- |
| 1 | PARAGUAY | 46.2 | 46.5 | 46.3 | 46.5 | 46.0 |
| 2 | PANAMA | 49.2 | 50.7 | 51.2 | 51.3 | 51.5 |
| 3 | BRAZIL | 53.9 | 58.4 | 58.9 | 59.6 | 61.7 |
| 4 | BOLIVIA | 42.2 | 42.2 | 43.2 | 42.7 | 42.0 |
| 5 | HONDURAS | 52.1 | 56.6 | 57.6 | - | 59.6 |
| 6 | DOMINICAN | 43.7 | 43.4 | 43.5 | 43.4 | 43.8 |
| 7 | CHILE | 46.0 | 46.9 | - | 47.0 | 48.3 |
| 8 | MEXICO | 45.4 | - | 45.8 | 45.7 | - |
| 9 | COLUMBIA | 50.4 | 51.3 | 51.4 | 51.7 | 52.8 |

**Table 2.** Data for the Analysis.

| S/N | Country Name | Gini Index | 10% Lowest Income | 10% Highest Income | S.F Index | D.I | Theil Index |
| --- | --- | --- | --- | --- | --- | --- | --- |
| 1 | Argentina | 41.4 | 1.8 | 29.9 | 16.61 | 0.58 | 0.312583 |
| 2 | Bolivia | 42.2 | 1.5 | 30.4 | 20.27 | 0.31 | 0.474293 |
| 3 | Brazil | 53.9 | 1.0 | 42.5 | 42.50 | 0.22 | 0.528996 |
| 4 | Chile | 46.0 | 1.8 | 37.1 | 20.61 | 0.44 | 0.529734 |
| 5 | Colombia | 50.4 | 1.4 | 39.7 | 28.36 | 0.27 | 0.569899 |
| 6 | Costa Rica | 48.0 | 1.5 | 36.3 | 24.20 | 0.31 | 0.429539 |
| 7 | Dominican | 43.7 | 2.3 | 35.2 | 15.30 | 0.37 | 0.429447 |
| 8 | Ecuador | 45.4 | 1.6 | 34.4 | 21.50 | 0.38 | 0.404237 |
| 9 | El Salvador | 38.6 | 2.4 | 29.4 | 12.25 | 0.58 | 0.327834 |
| 10 | Guatemala | 48.3 | 1.7 | 38.0 | 22.35 | 0.26 | 0.509287 |
| 11 | Honduras | 52.1 | 0.9 | 39.1 | 43.44 | 0.16 | 0.476235 |
| 12 | Mexico | 45.4 | 2.0 | 36.4 | 18.20 | 0.30 | 0.511219 |
| 13 | Nicaragua | 46.2 | 2.0 | 37.2 | 18.6 | 0.46 | 0.479821 |
| 14 | Panama | 49.2 | 1.2 | 37.1 | 30.92 | 0.24 | 0.491384 |
| 15 | Paraguay | 46.2 | 1.7 | 35.9 | 21.12 | 0.31 | 0.618109 |
| 16 | Peru | 42.8 | 1.8 | 32.1 | 17.83 | 0.29 | 0.355739 |
| 17 | Uruguay | 39.7 | 2.3 | 29.7 | 12.91 | 0.71 | 0.307564 |

### 3.2 Epidemiologic Variables

The variables from epidemiology were chosen during the exponential phase of the first and second wave of the pandemic. Daily new cases observed during the first 100 days was used to calculate the exponential slope for first and second wave, opposite autocorrelation slope were averaged on six days for the first and second wave. The maximum R_o_ was collated from [16] while observing this value during the first and second waves of countries considered. We also collated from [16] the deterministic R_o_ for the first and second wave of the pandemic taking six days as length of contagiousness period.

So, in summary we have six variables from epidemiology which are: first wave maximum R_o_, second wave maximum R_o_, first wave deterministic R_o_, second wave deterministic R_o_, opposite initial autocorrelation slope averaged on six days for both first and second wave of the daily new cases for developed and developing countries. All epidemiologic variables values were taken from the Appendix in [16].

In this present study, we validated our results by performing cross validation and also training 80% of the data and training 30%.

## 4 The Relationship Between Theil and Gini Index

### 4.1 Mathematical Approach

We first show the relationship between Theil and Gini index mathematically:

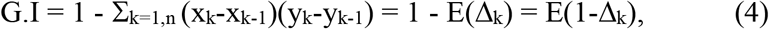

where x_k_ (respectively y_k_) denotes the k^th^ cumulative part of the population (respectively income). If we choose the population increments equal to 1/n and if E(Δ_k_) represents the expectation for the distribution d_k_ = x_k_ - x_k-1_ of the increment Δ_k_ = y_k_-y_k-1_ (see [17]), we have for the Theil index applied to the percentage y_k_ of the total income relative to a percentage x_k_ of the total population (see [18]):

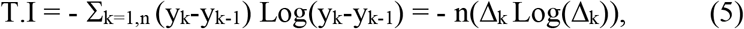

If the first increment of y is close to 1 (which corresponds to a square shaped Lorenz curve, i.e., closed to the down and left border of the income/population square, or to a high Gini index) (cf. Figure 1), then -Log(Δ_1_)∼-1+Δ_1_, and we have:

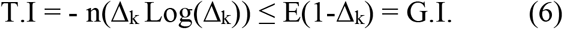

the equality being available only if the Lorenz curve presents a perfect top left right-angle shape.

**Figure 1.**
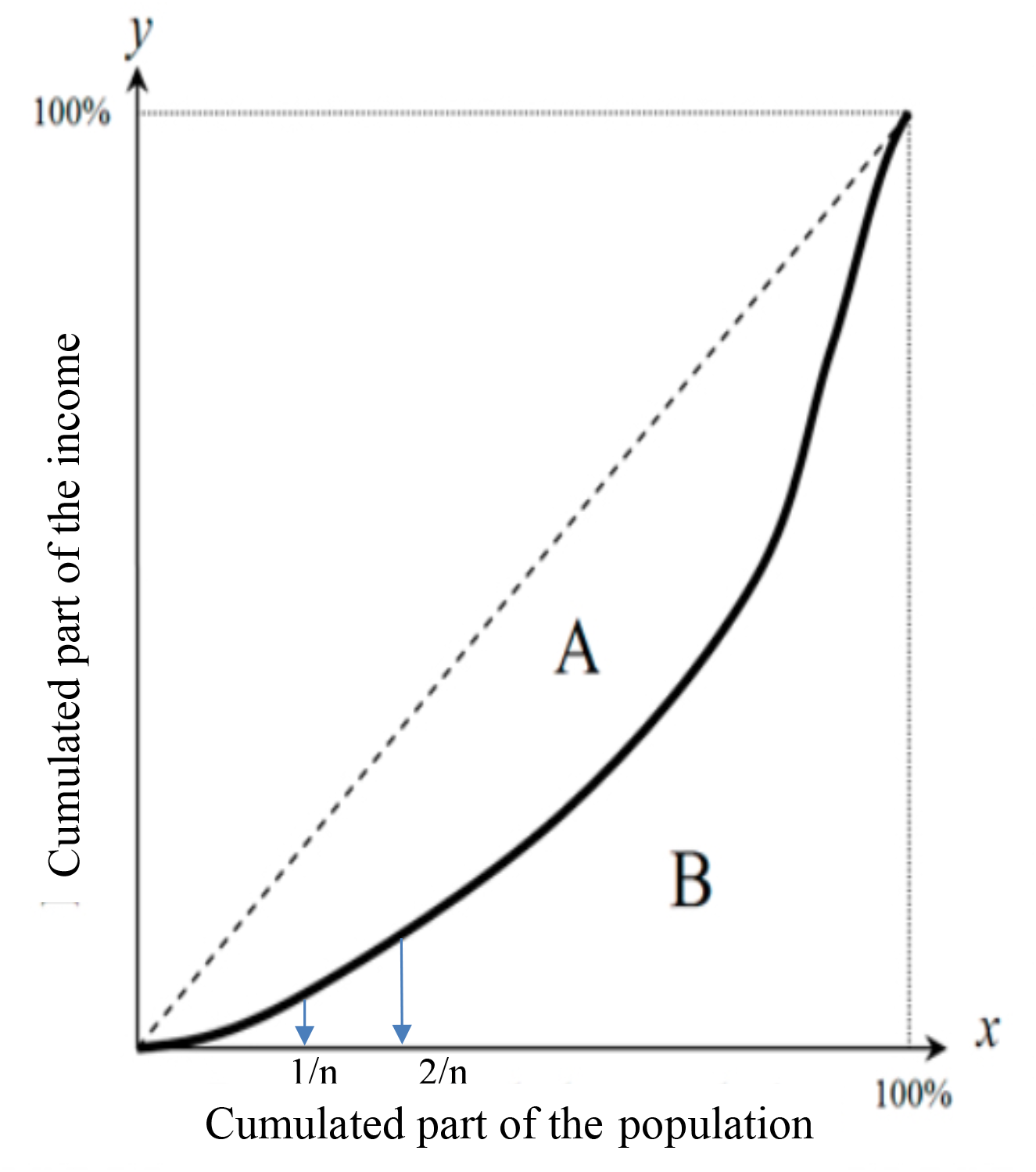
Lorenz curve showing the cumulated part of income vs cumulated part of a population having this cumulated income.

### 4.2 Statistical Approach

#### 4.2.1 Correlation

We correlated both Theil and Gini Indices with all epidemiologic, demographic and socio-economic variables and as it can be seen in Figure 2, Theil and Gini Indices are highly positively correlated with coefficient 0.7.

**Figure 2.**
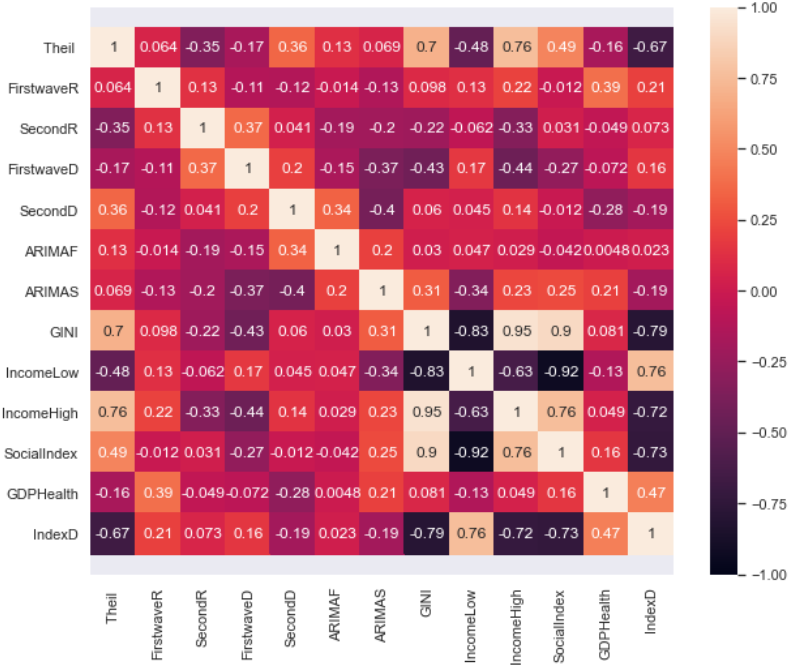
Heat map for the correlations between all variables.

### 4.2.2 Regression Analysis Between Theil and Gini Index

Linear regression models use some historic data of independent and dependent variables and consider a linear relationship between both while polynomial regression models use a similar approach but the dependent variable is modeled as a degree n (n=2 in the present study) polynomial in x.

Linear regression model is given as:

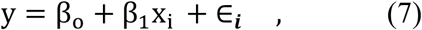

where β_1_ are the weights, β_0_ is the intercept and ∈ is the random error term. The above equation is the linear equation that needs to be obtained with the minimum error. Polynomial regression of order 2 is given below:

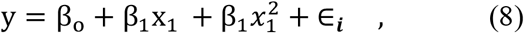

We present the visualization of the results using this approach in Figure 3.

**Figure 3.**
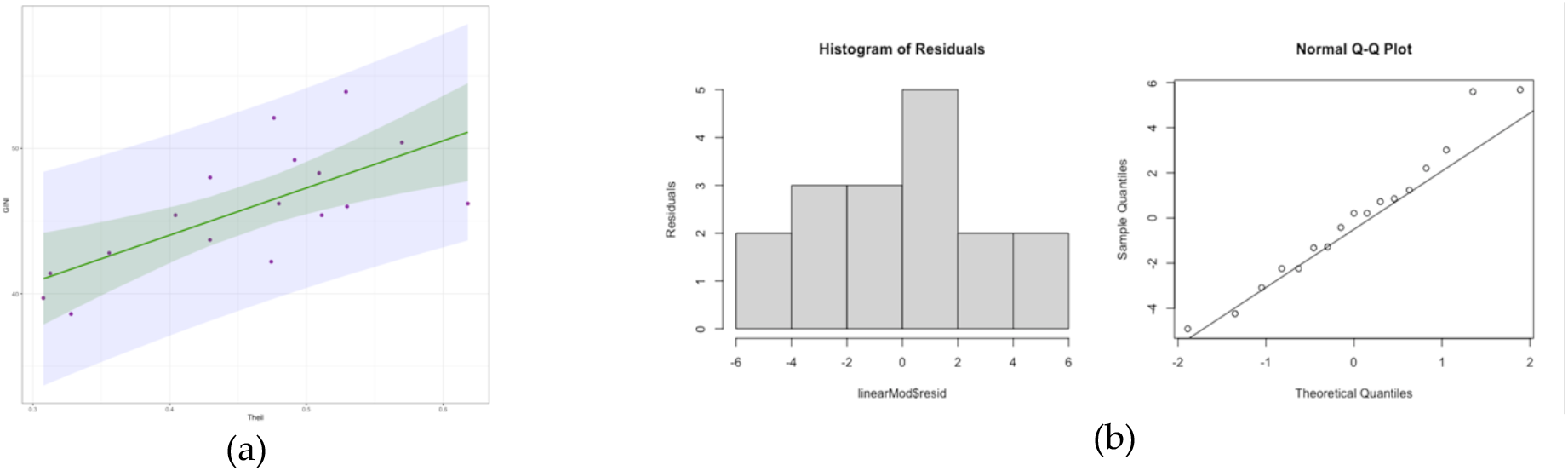
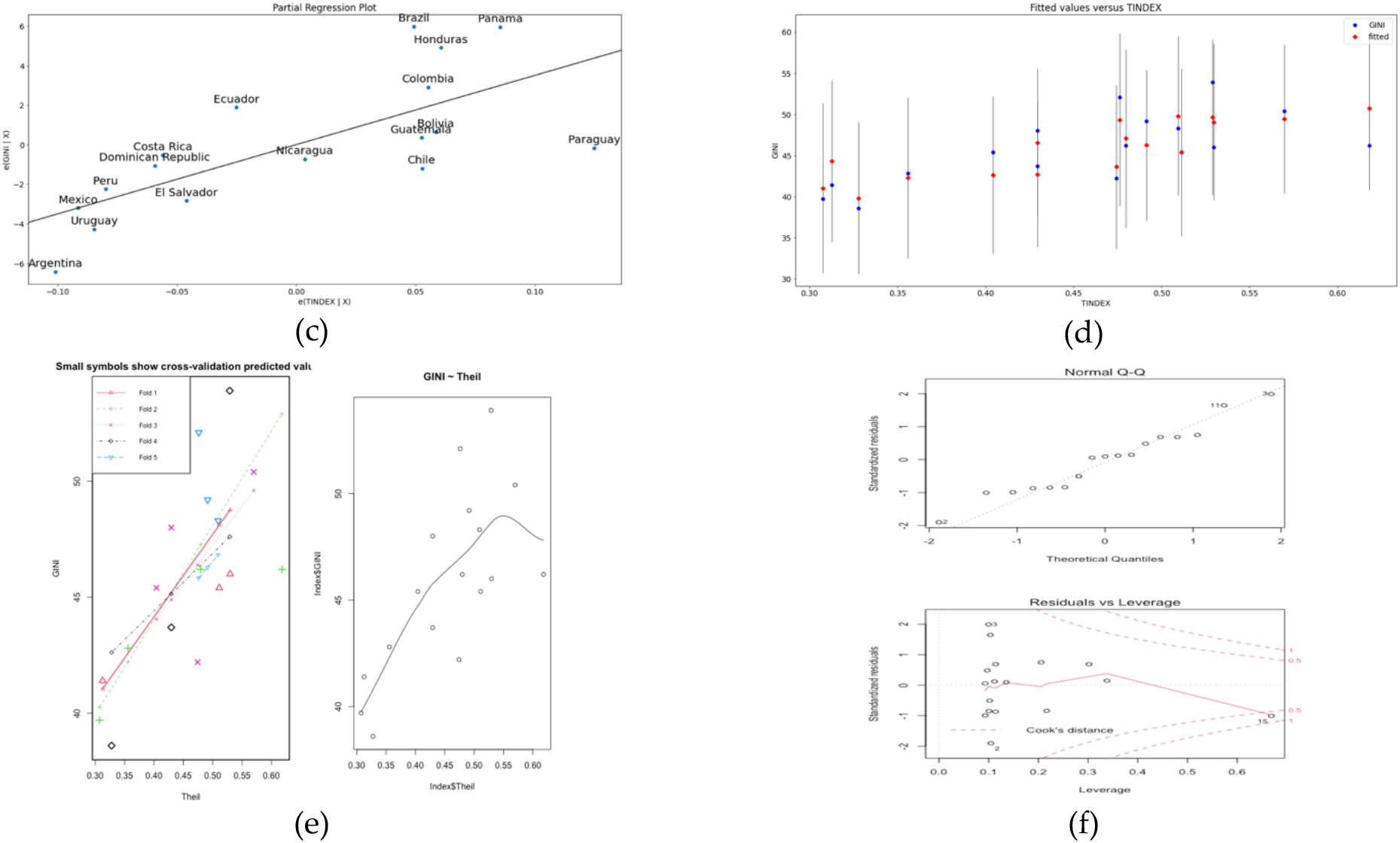
(a) Linear regression line (in green) with the confidence interval (in grey). (b) Residual plot for linear regression. (c) Partial regression plot. (d) Fit plot. (e) On left hand side is the cross-validation plot for the linear regression and on right-hand side is the polynomial regression plot. (f) Residual plots for polynomial regression.

For the linear regression as shown in Figure 3a, the intercept is 31.03, p-value is 0.0181, R-squared is 0.4881, residual standard error is 3.116 and all coefficients are significant with p < 0.05 for both the train and test data for linear and polynomial regression. The median of the residual plot in Figures 3b and 3f are 0.2111 and 0.2566 respectively for both linear and polynomial regression which is close to zero. The normality of the residual was tested using Jarques Bera and Durbin-Watson tests which gave a high p-value and we fail to reject the null hypothesis that the skewness and kurtosis of the residuals are statistically equal to zero. In order to know the performance of the linear regression model we trained 80% of the data and tested 20% of the data and also did cross validation to be sure of the accuracy. The predicted and the observed values are very close to the result presented for the regression models used. For the linear model we present the cross-validation result in Figure 3e whose average mean square error for the 5 portion folds is 11.72794. We observed correlation between the tested and the predicted values has high correlation accuracy (R-squared = 0.97). The test set p-value is 0.02 with residual standard error of 3.528. For polynomial regression of order 2, the train set has the following results: R-squared = 0.6, p-value = 0.002 and residual standard error = 2.935. The test set has the following results: R-squared = 0.99, p-value = 0.008 and residual standard error = 0.5639.

#### 4.2.3 Neural Network for Theil and Gini Index

We used the *neuralnet* package in R in order to visualize the weights of the network and the bias between Theil and Gini index and as it can be seen in Figure 4, the weights are good with low bias. We also predicted some of the data and the prediction score is 0.98.

**Figure 4.**
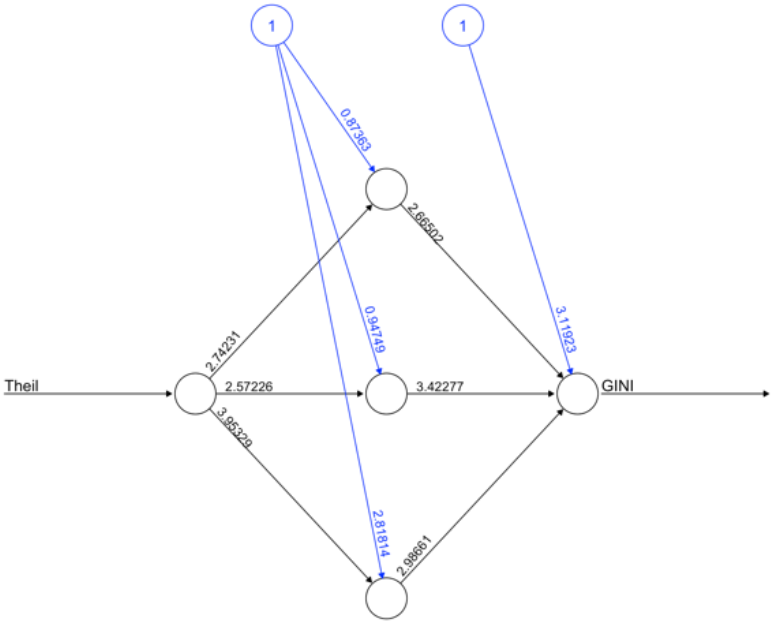
Neural network visualization.

### 4.2.4 Multivariate Analysis for Gini and Theil Index Alongside Other Socio-economic Variables and Epidemiologic Variables

Figure 5 corresponds to the ordinary multivariate least square methods with R-squared = 0.674. Figure 5a shows Paraguay as outliers not fitting the data, Figure 5b normalizes all countries and doesn’t point any country in the plot.

**Figure 5.**
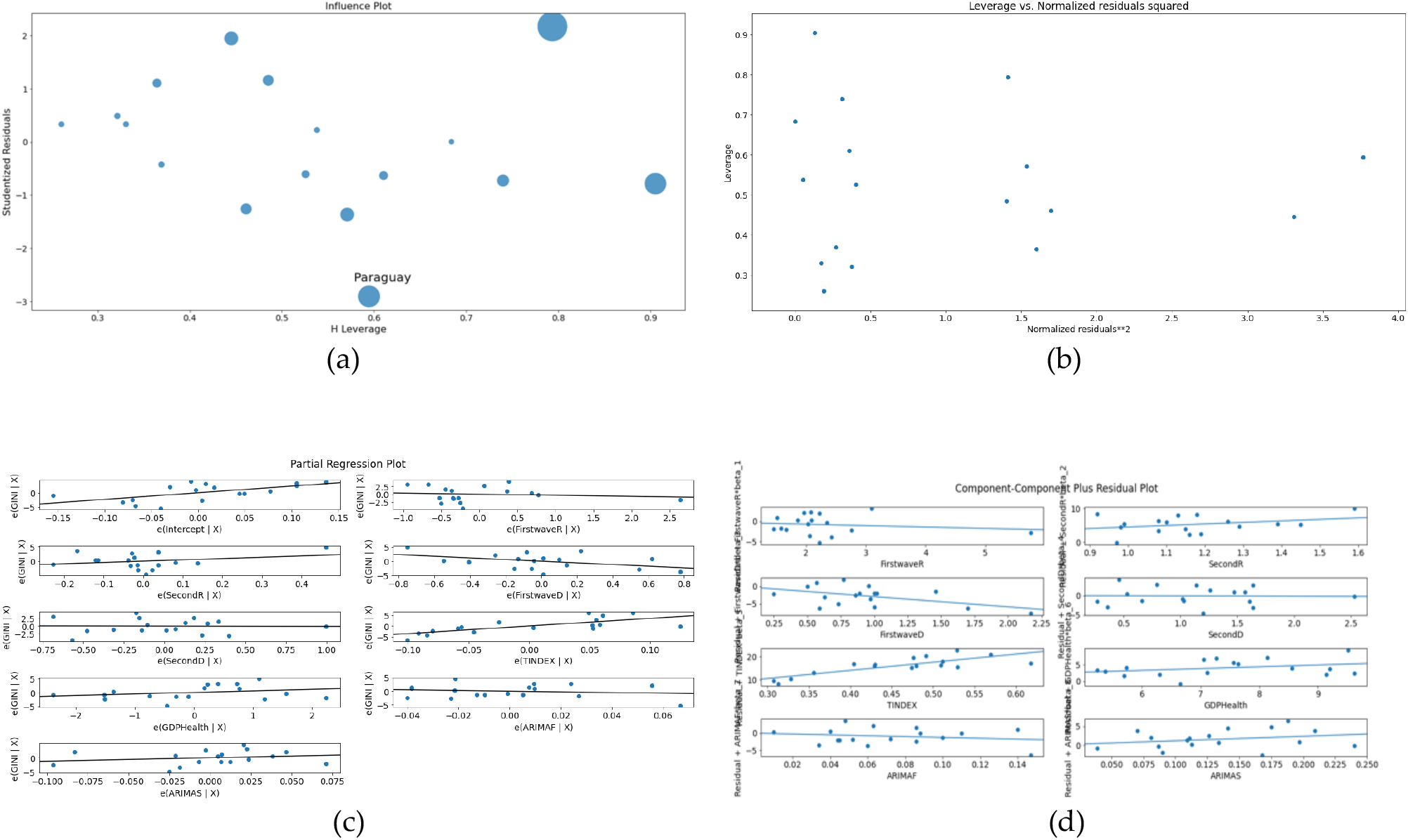
(a) Influence plot. (b) Leverage vs Normalized residuals squared plot. (c) Partial regression plot. (d) Component-Component plus residual plot.

### 4.2.5 Prediction of Gini Index Using MLP Regressor, Linear, Lasso and Ridge Regression

In this section we used cross validation method to choose the best parameter α for the modelling as shown in Figure 6c. For ridge regression, α = 0.142 with mean square error of 1.36 and α = 0.368 for lasso regression with mean square error = 5.10. For Figure 6e, the training score = 1.000 and the test score = 0.641, for Figure 6f training score = 0.99 2 and the test score = 0.497, for Figure 6g training score = 0.99 and the test score = 0.406 and for Figure 6h training score = 0.984 and test score = -0.077. It is evident from these results that linear regression best predicts Gini index with the highest test score and predicted values are very close to each other as presented in Table 1. Also, we observed the same pattern of prediction in Figures 6e to 6h showing that the all methods used in this section have the same predictive behavior.

**Figure 6.**
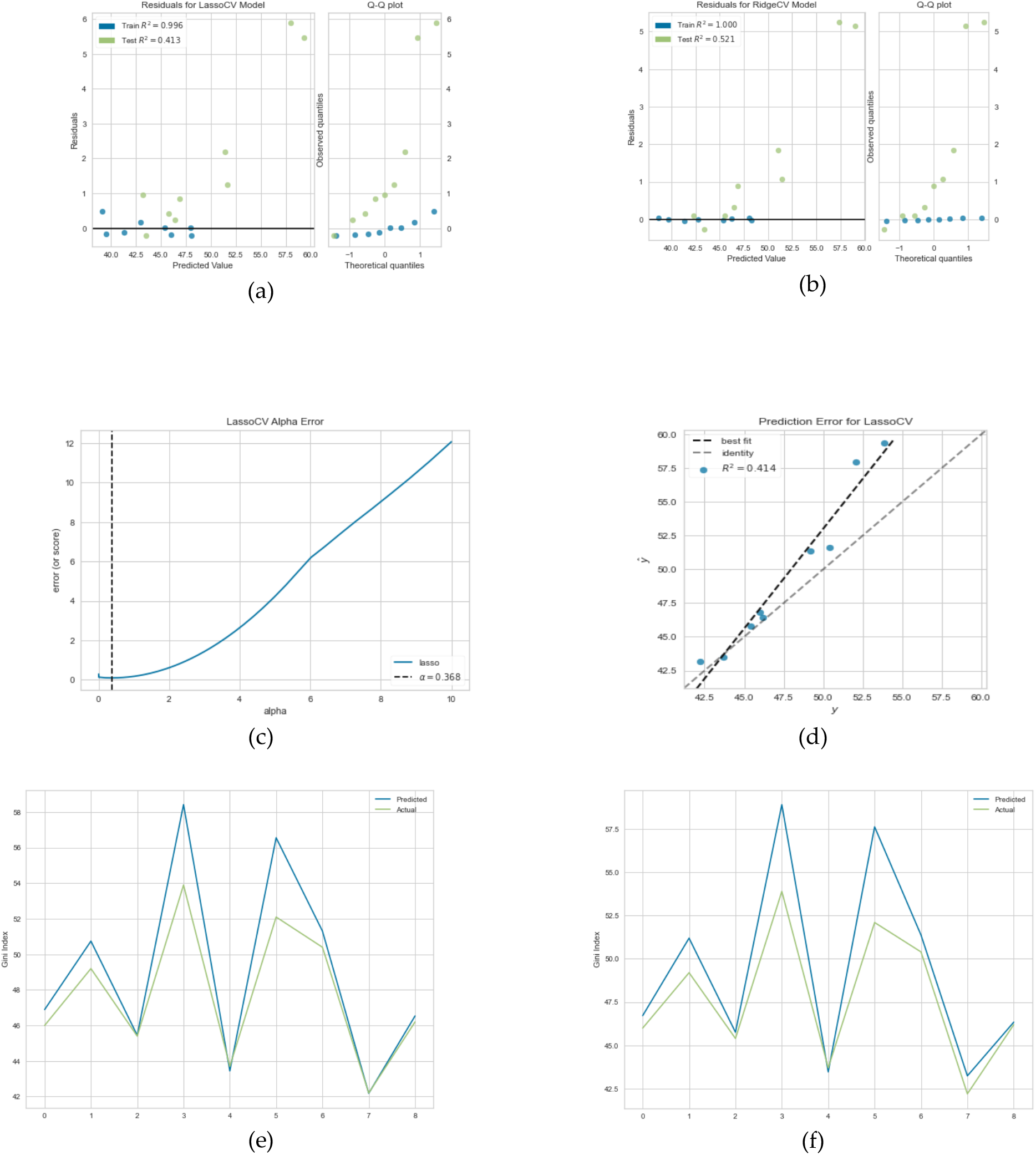
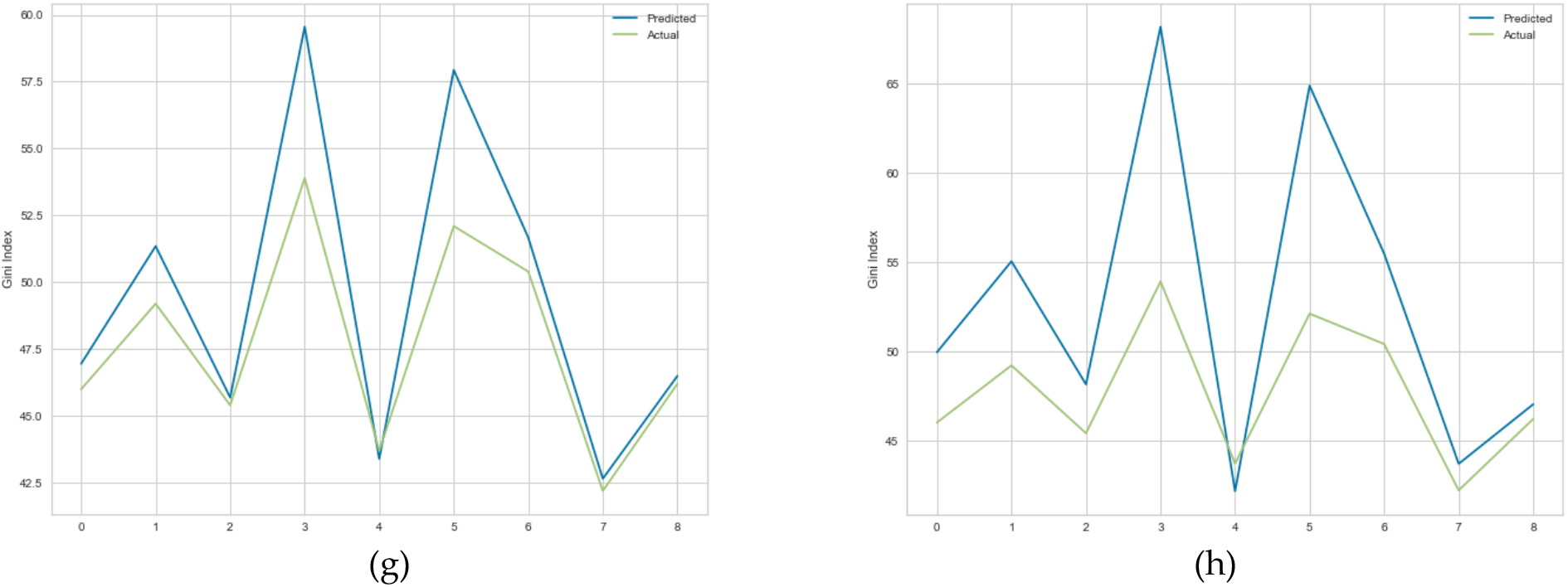
(a) Residual plot for lasso regression. (b) Residual plot for ridge regression. (c) Lasso regression cross validation error. (d) Prediction error for lasso regression. (e) Linear regression prediction plot. (e) Linear regression prediction plot. (f) Lasso regression prediction plot. (g) Ridge regression prediction plot. (f) MLP regression prediction plot.

#### 4.2.6 Clustering Analysis for Gini and Theil Index Alongside Other Socio-economic Variables and Epidemiologic Variables

In Figure 7c, the first cluster has 14 countries and the second has 3 countries which are Uruguay and El Salvador on same hierarchy while Argentina is on another hierarchy. We only show the clusters dendrogram for the first cluster. In Figure 7f, Gini index has the highest positive correlation of 0.44 and Theil index has the value 0.34 in PC 1.

**Figure 7.**
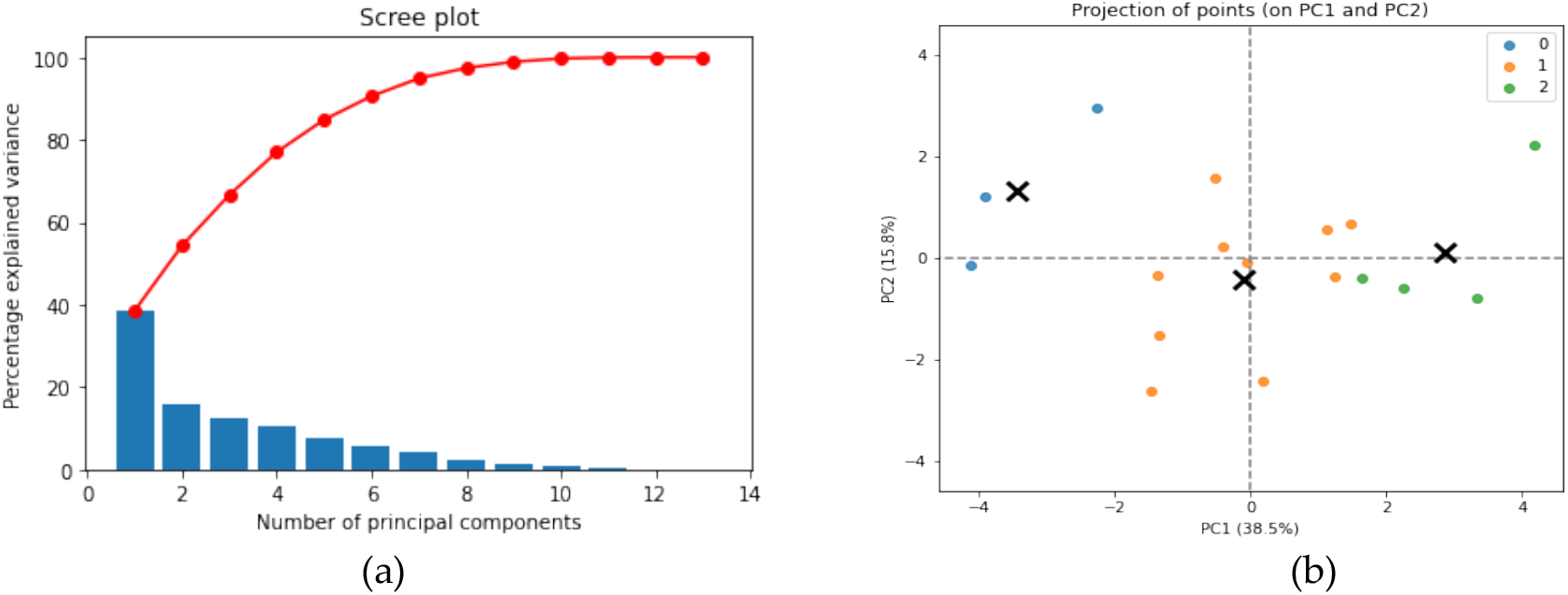
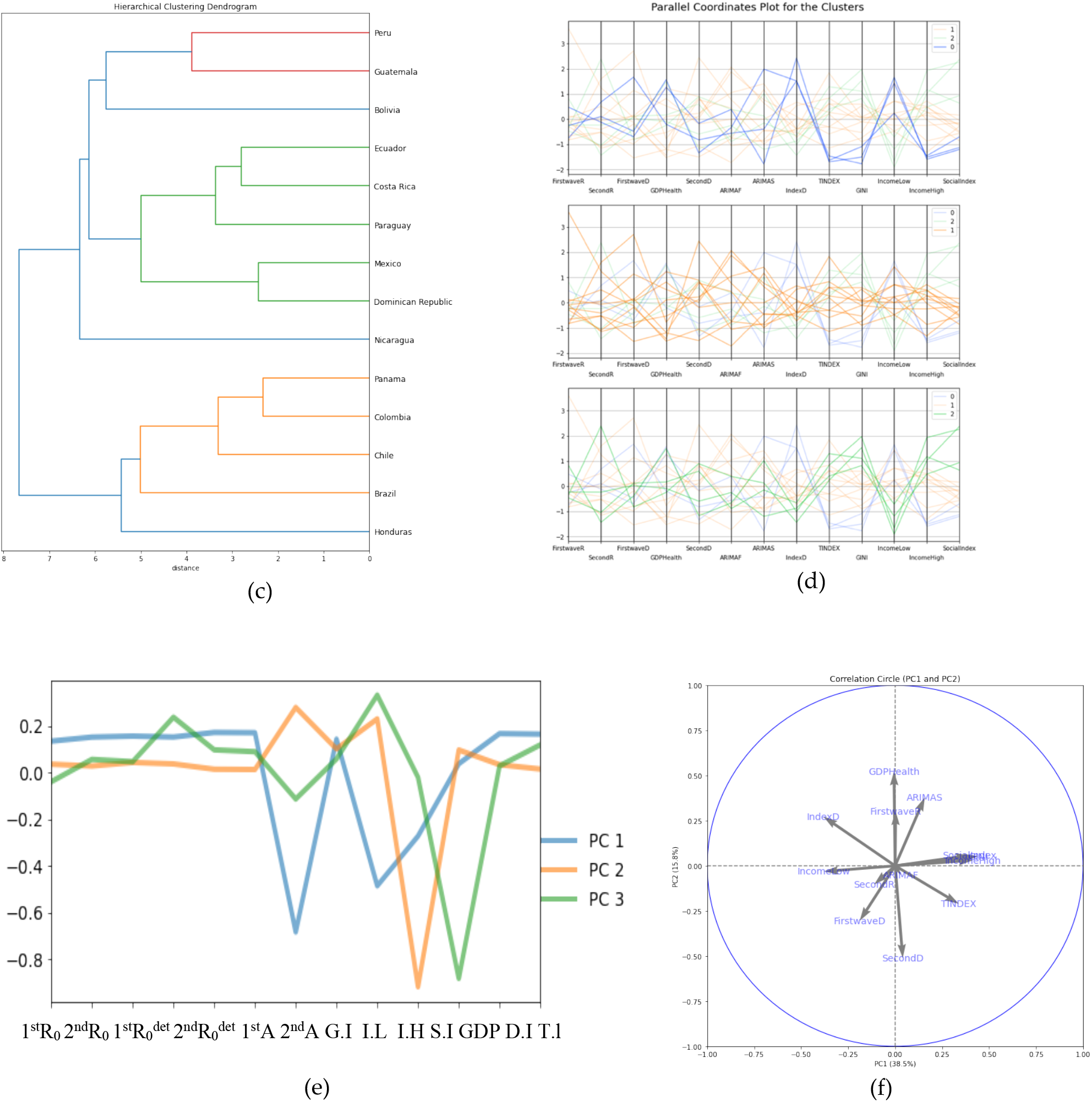
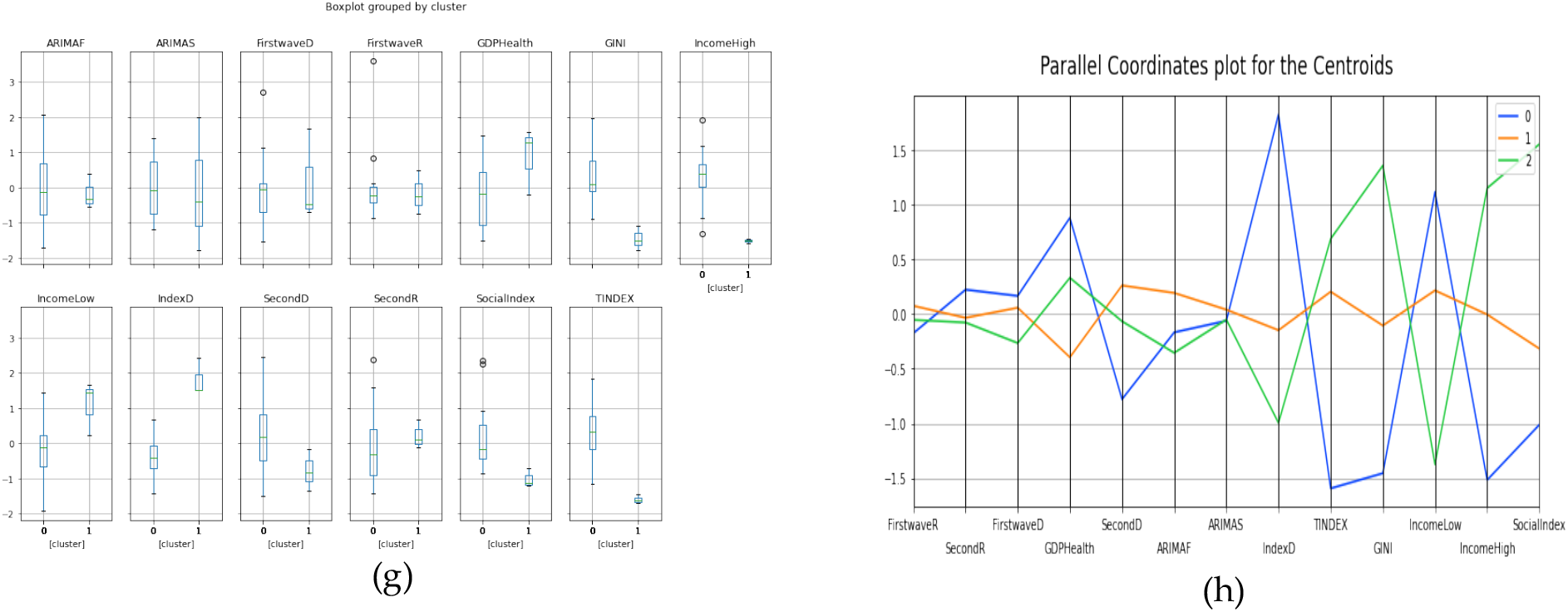
(a) Scree plot. (b) Plot for projection of points for PC1 and PC2. (c) Hierarchy clustering dendrogram. (d) Parallel coordinates plot for the clusters. (e) & (f) PC’s visualization. (g) Box plot for the clusters. (h) Parallel coordinate plot for the centroids.

## 5 Application of the Methods to OECD Countries, Africa Countries, Developed and Developing countries

### 5.1 Developed and Developing Countries

#### 5.1.1 Regression and Multivariate Analysis for Socio-economic Variables and Epidemiologic Variables

In Figures 8c and 8d we modeled the dependent variable as a degree n (n=6 in the present study) polynomial in x, an extension of Equation 8. The Figure 8 present regression analyses with the following parameters: Figure 8a: LinregressResult (slope = 0.11463663009107196, intercept = -0.0037118697103040027, rvalue = 0.43871 57758684147, pvalue = 0.0032517683682962654, stderr = 0.03667137141150123, R-squared = 0.192472 and RMSE = 0.1044724946057671.)

**Figure 8.**
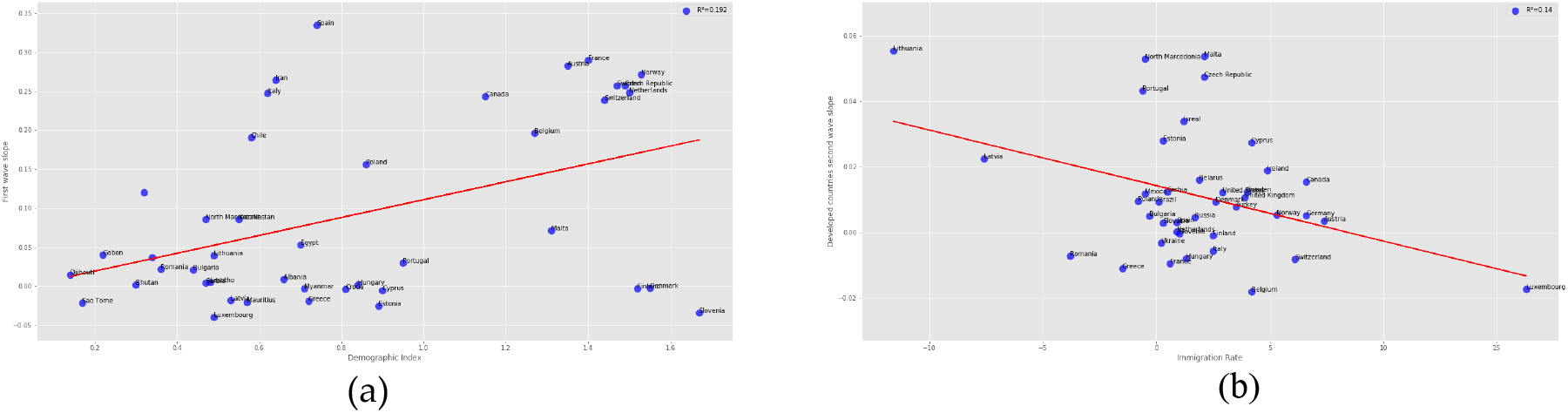
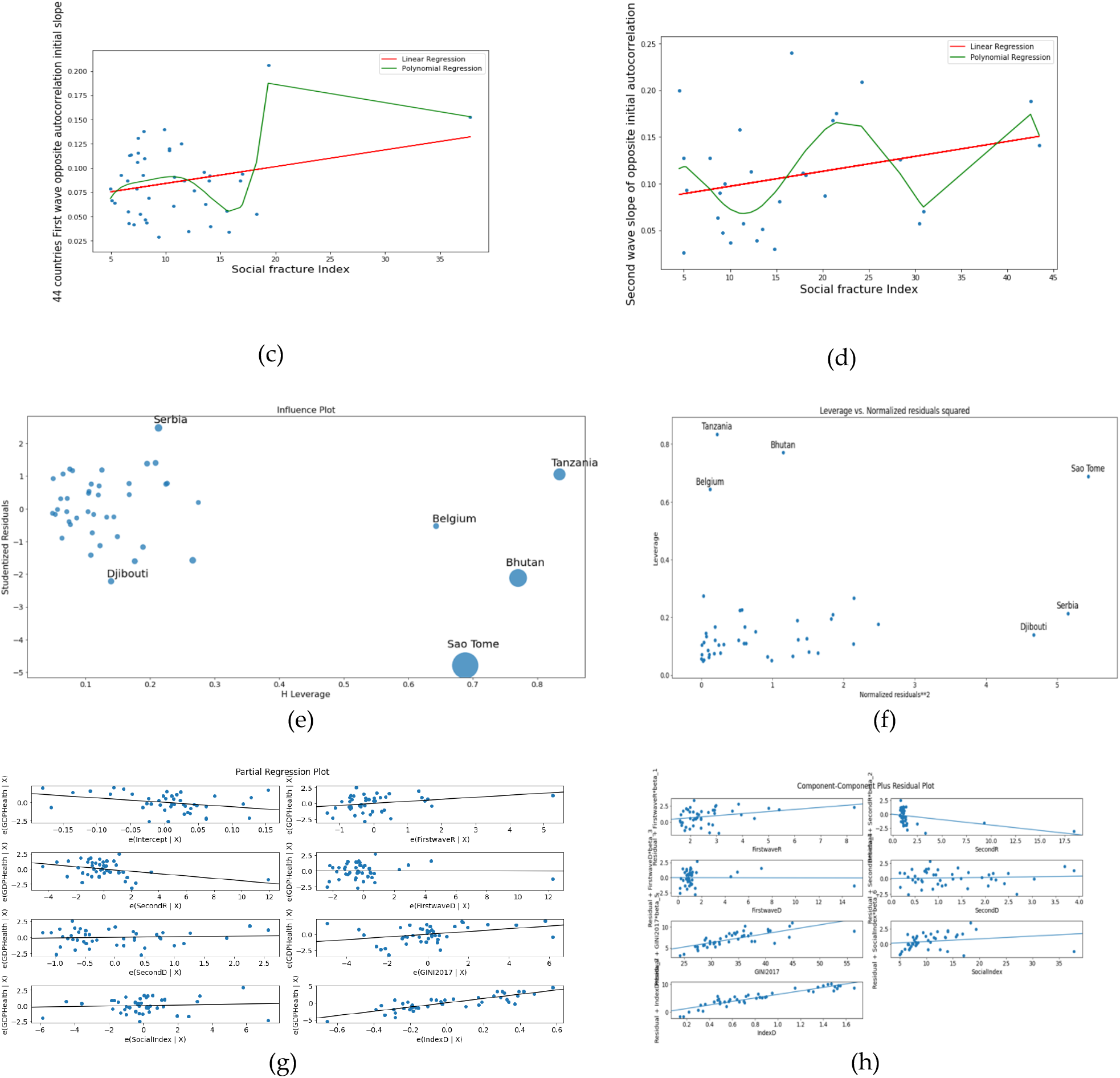
Linear regression plots for (a) first wave slope vs. demographic index for developed and developing countries, (b) second wave slop vs. immigration rate for developed countries, (c) opposite of the initial auto-correlation slope for first wave vs. social fracture index and (d) opposite of the initial autocorrelation slope for second wave vs. social fracture index. (e) Influence plot. (f) Leverage vs Normalized residuals squared plot. (g) Partial regression plot. (h) Component-Component plus residual plot.

Figure 8b: LinregressResult (slope = -0.002547609589041096, intercept = 0.07888755616438356, rvalue = -0.32728 86357381478, pvalue = 0.03672803730354382, stderr = 0.0011777868896598461,

R-squared = 0.107118 and RMSE = 0.03065537183298402)

Figure 8c: LinregressResult (slope = 0.0017309145398433248, intercept = 0.06695128460299407, rvalue = 0.26367 5660748951, pvalue = 0.08754941979369255, stderr = 0.0009889311191763849,

R-squared = 0.069525 and RMSE = 0.0354860744891158), R-squared for order Six Polynomial Regression = 0.3 and RMSE for Polynomial Regression of order six = 0.04060485094256808.

Figure 8d: LinregressResult (slope = 0.0015999465132904799, intercept = 0.0810899892250729, rvalue = 0.286126 6574746827, pvalue = 0.13239511534872409, stderr = 0.001031140187045727, R-squared = 0.081868 and RMSE = 0.05492215494302141), RMSE for Polynomial Regression of order six = 0.07286590609946085 and R-squared for order Six Polynomial Regression = 0.35.

Figures 8e to 8h correspond to the ordinary multivariate least square method with R-squared = 0.76. Figure 5a shows some developing countries as outliers while Belgium is the only developed country which do es not fit the data.

#### 5.1.2 Prediction of Percentage GDP Health Expenditure

In this section we used cross validation method to choose the best parameter α for the modelling as shown in Figure 9c. For ridge regression, α = 0.012 with mean square error of 2.32 and α = 0.029 for lasso regression with mean square error = 2.21. For Figure 9e, the training score = 0.983 and the test score = 0.607, for Figure 9f training score = 0.170 and the test score = 0.021, for Figure 9g training score = 0.854 and the test score = 0.115 and for Figure 9h training score = 0.980 and test score = -2.386.

**Figure 9.**
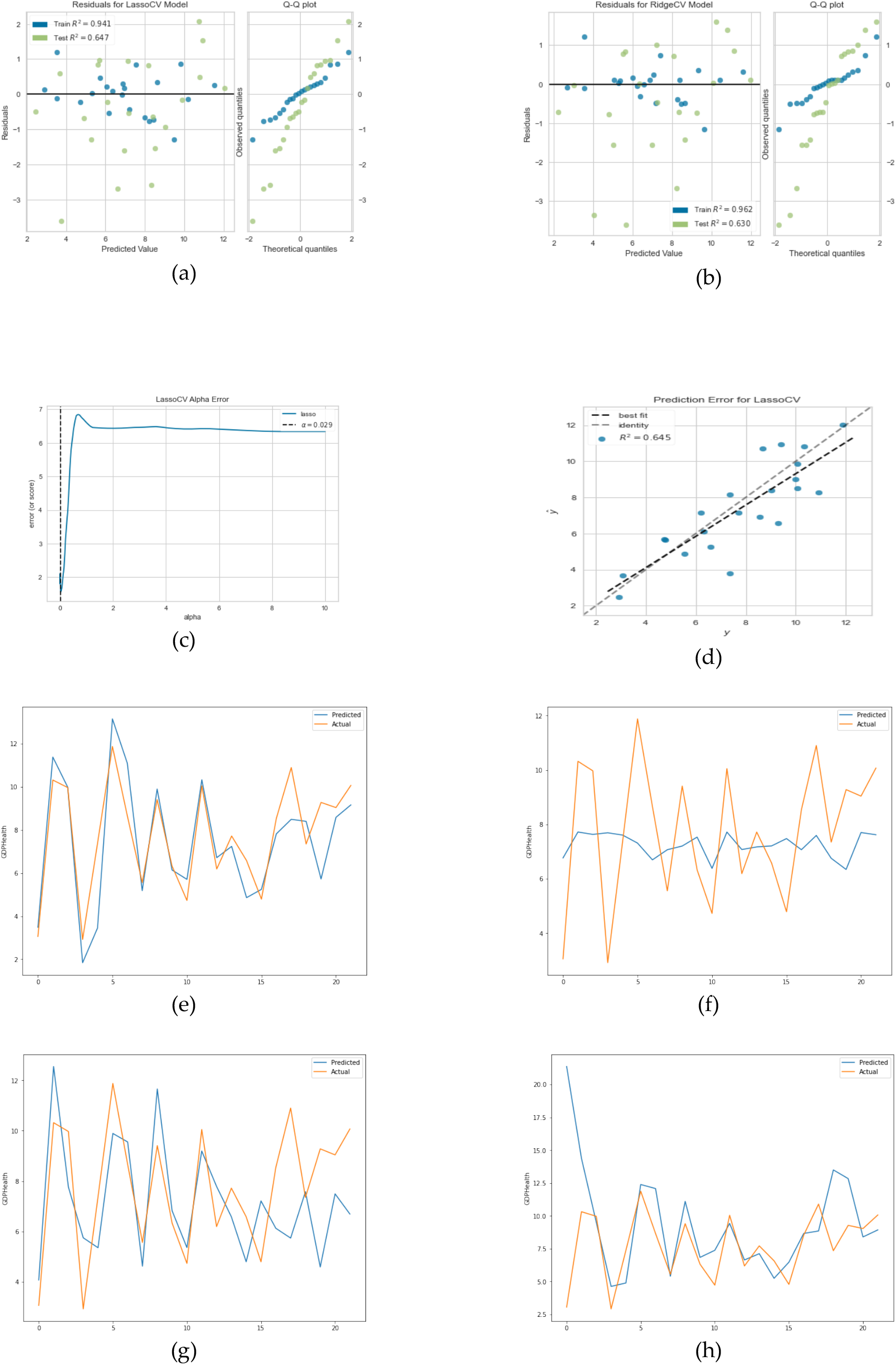
(a) Residual plot for lasso regression. (b) Residual plot for ridge regression. (c) Lasso regression cross validation error. (d) Prediction error for lasso regression. (e) Linear regression prediction plot. (e) Linear regression prediction plot. (f) Lasso regression prediction plot. (g) Ridge regression prediction plot. (h) MLP regressor prediction plot.

It is evident from the results that linear regression best predicts percentage of GDP devoted to health expenditure with the highest test score and predicted values are very close.

#### 5.1.3 Principal Component Analysis and Clustering Result

In Figures 10e and 10f, the first cluster has 15 countries, the second cluster 27 countries while the last cluster has 2 countries which are Tanzania and Mauritius. We only show the two clusters dendrograms with many countries. In Figure 10c, Gini index has the highest positive correlation of 0.52 and demographic index has highly negative correlation of -0.53 in PC 1 while first wave maximum R_o_ has highest positive correlation in PC 2, whose value equals 0.70.

**Figure 10.**
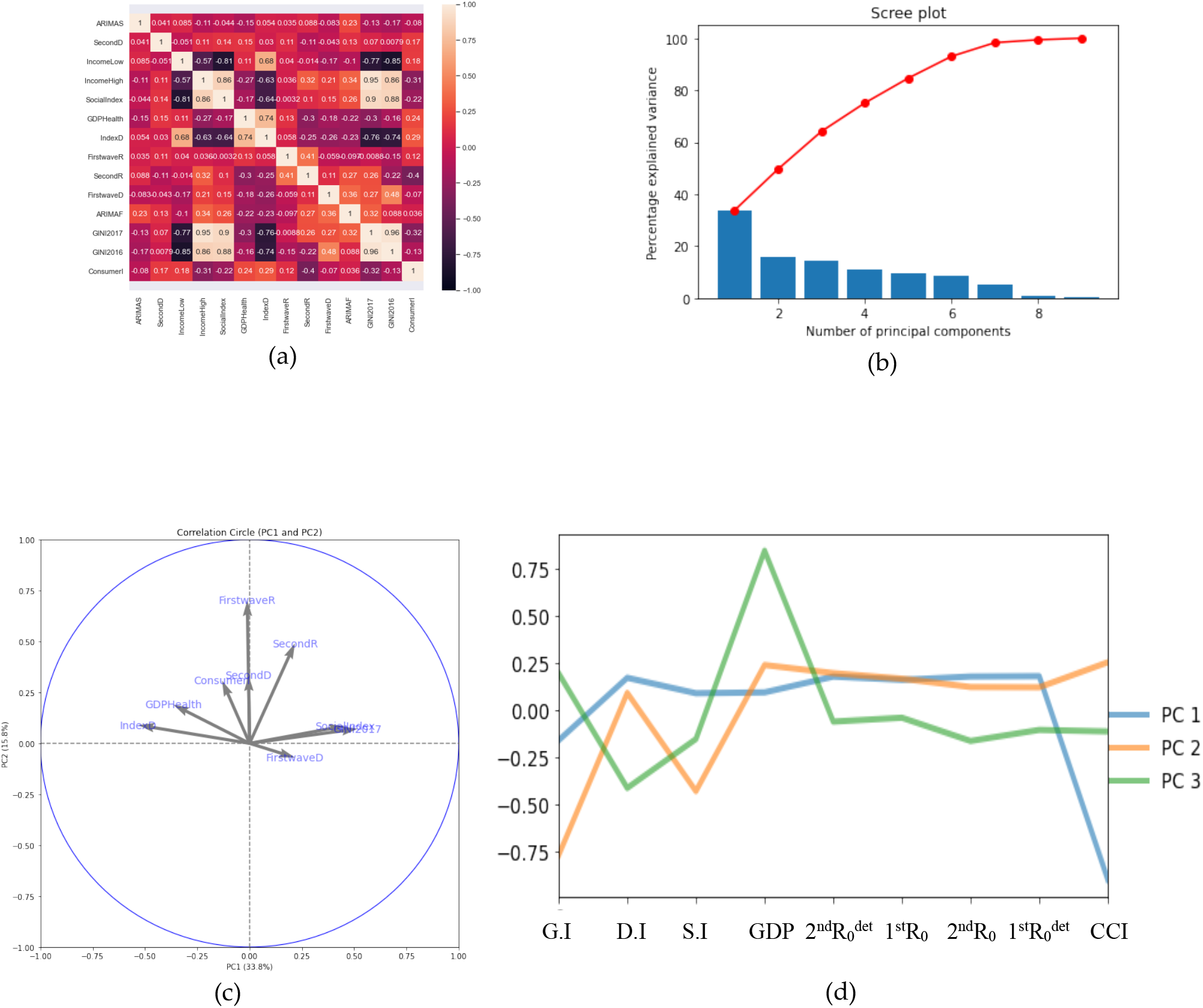
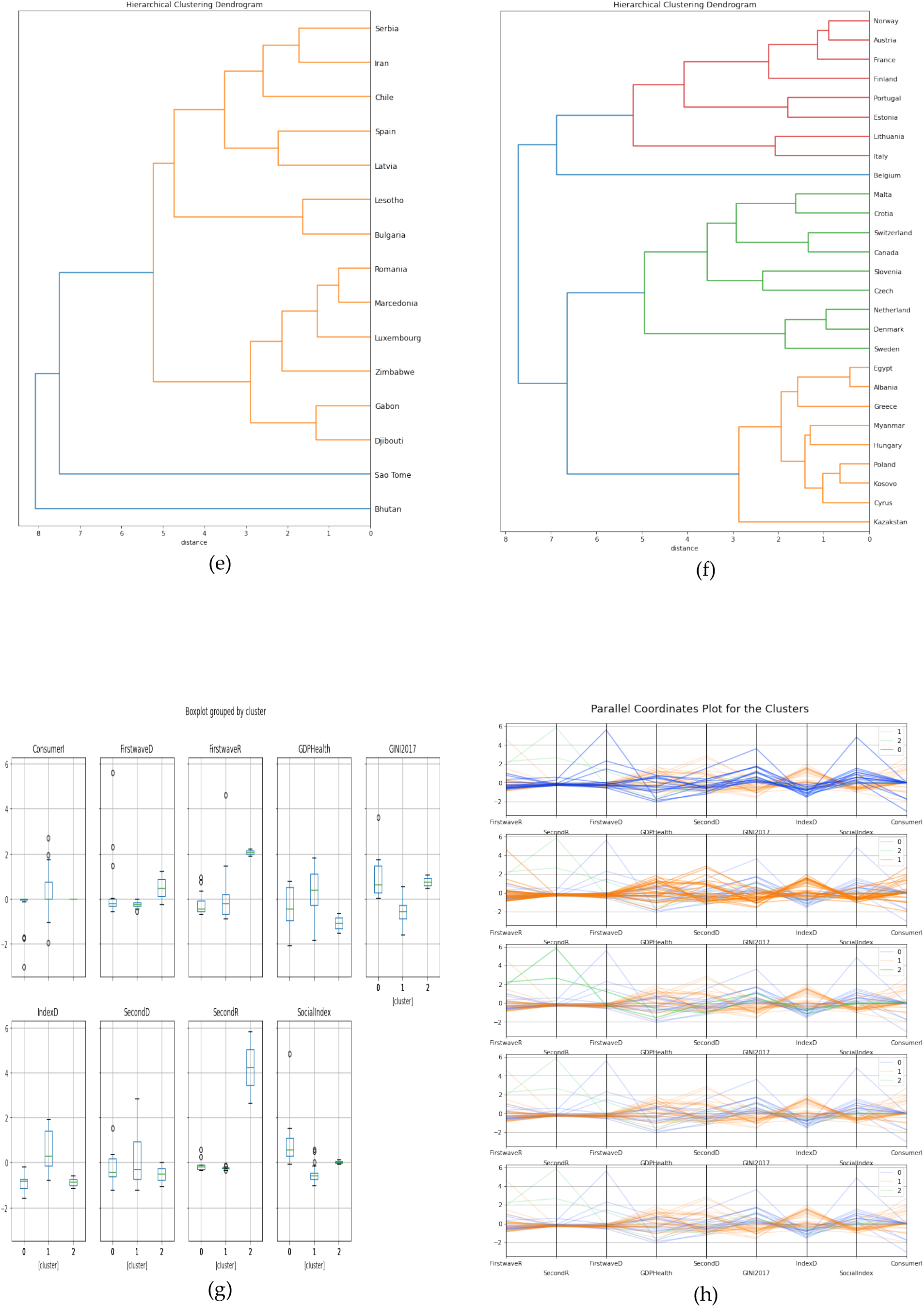
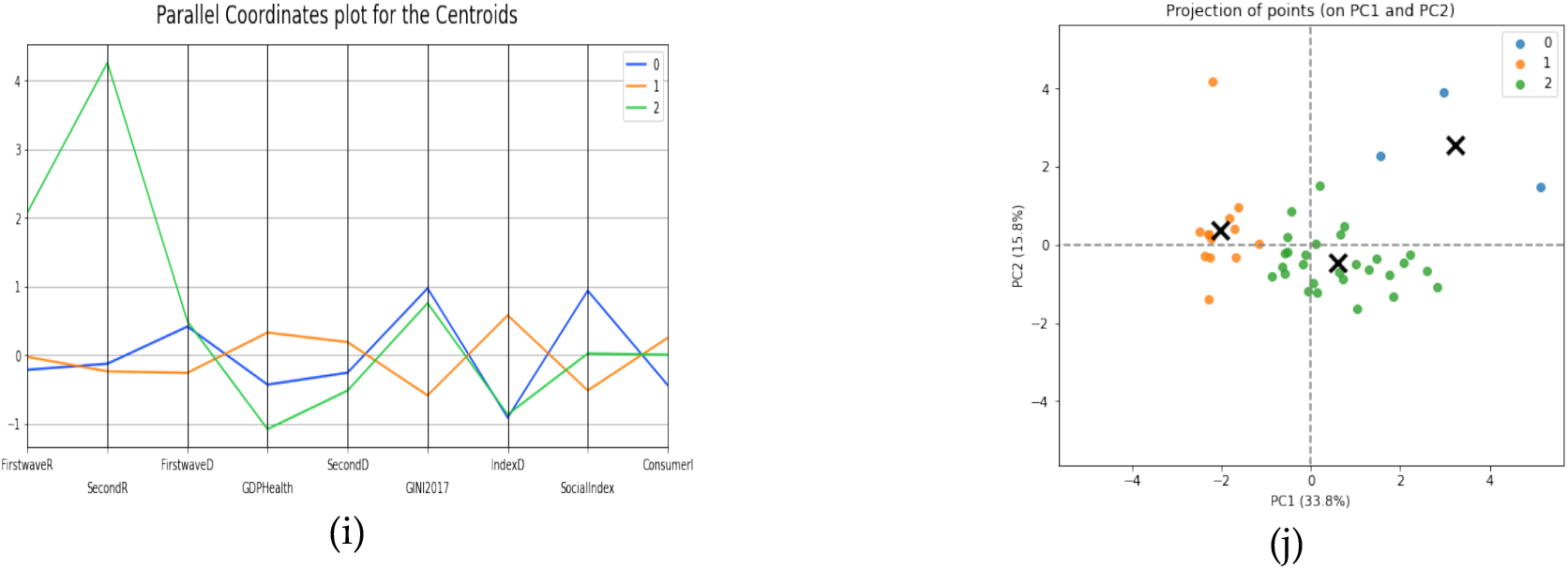
(a) Heatmap of the parameter’s correlations. (b) Scree plot. (c) & (d) PCs visualization. (e) & (f) Hierarchy clustering dendrogram. (g) Boxplot of the clusters. (h) Parallel coordinates plot for the clusters. (i) Parallel coordinates plot for the centroids. (j) Projection points for PC1 and PC2.

### 5.2 Africa Countries

### 5.2.1 Multivariate Analysis for Socio-economic Variables and Epidemiologic Variables

Figures 11a to 11d correspond to the ordinary multivariate least square method with R-squared = 0.60. Figure 11a shows Botswana and Tanzania as outliers not fitting the data.

**Figure 11.**
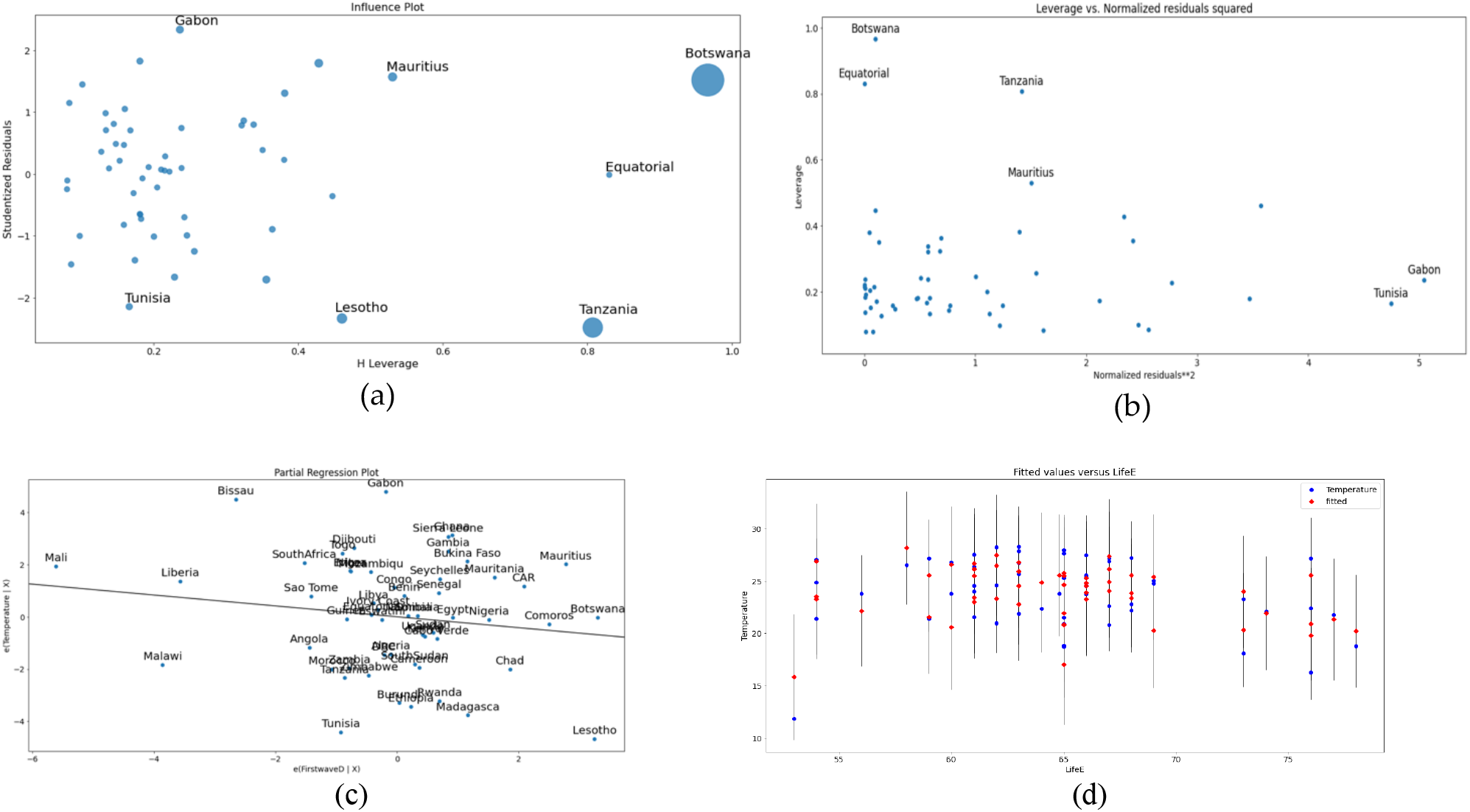
(a) Influence plot, (b) Leverage vs Normalized residuals squared plot, (c) Partial regression plot and (d)Fit plot.

#### 5.2.2 Prediction of Temperature

In this section we used cross validation method to choose the best parameter α for the modelling as shown in Figure 12c. For ridge regression, α = 1.005 with mean square error of 19.13 and for lasso regression α = 6.018 with mean square error = 16.93. For Figure 12e, the training score = 0.647 and the test score = -2.228, for Figure 12f training score = 0.316 and the test score = 0.154, for Figure 12g training score = 0.573 and the test score = -1.136 and for Figure 12h training score = -6.728 and test score = -4.714. It is evident from these results that lasso regression best predicts temperature with the highest test score and predicted values are close.

**Figure 12.**
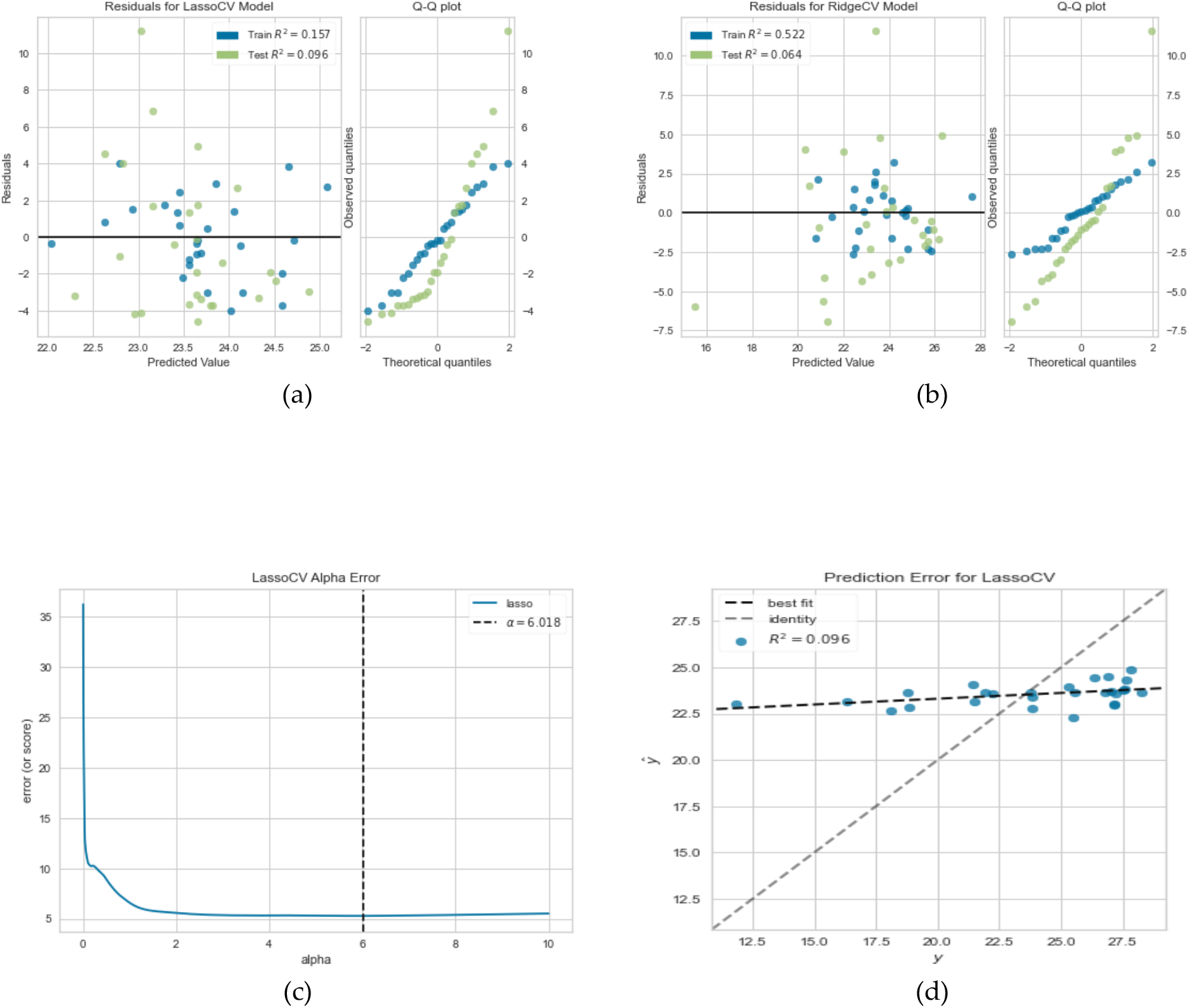
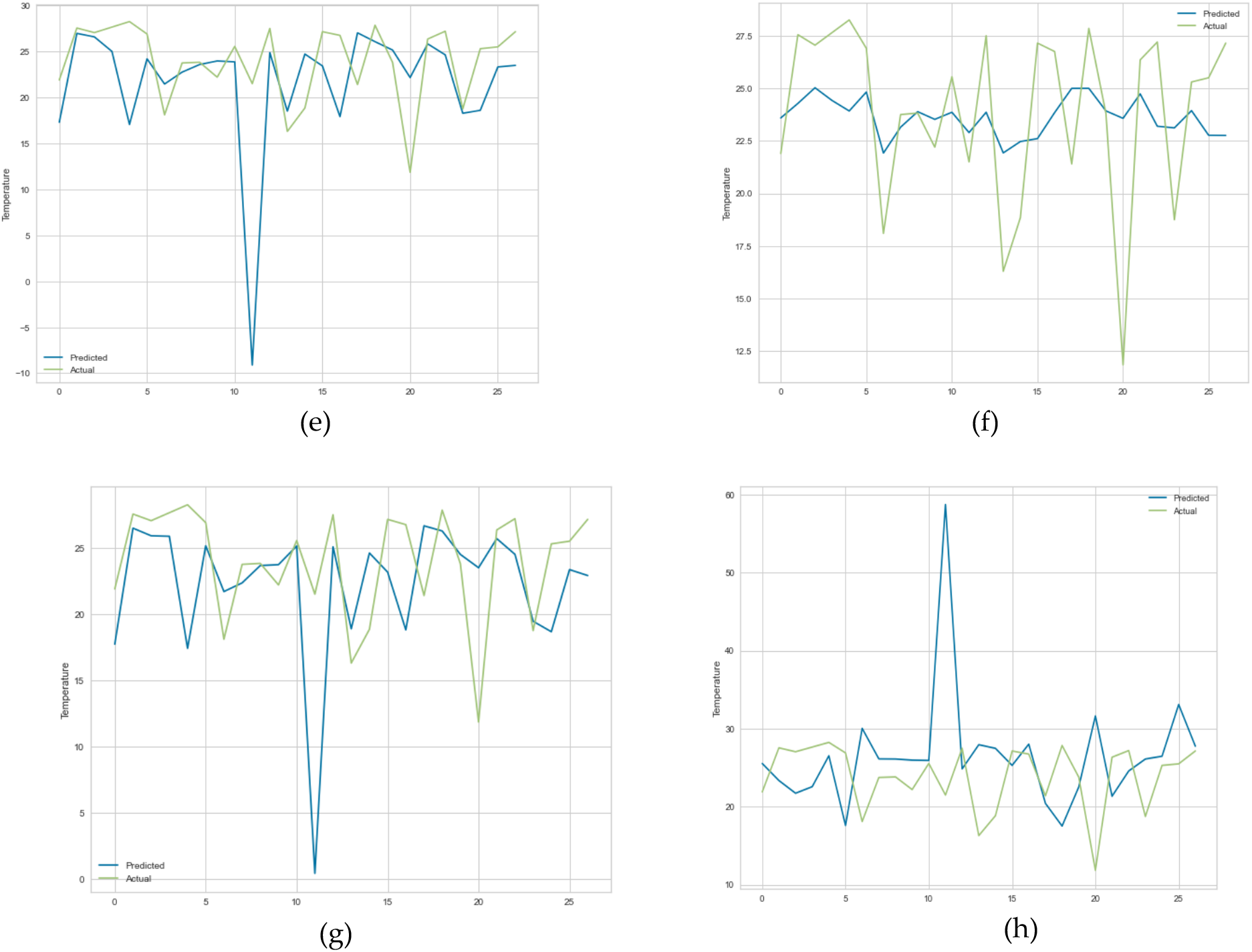
(a) Residual plot for lasso regression, (b) Residual plot for ridge regression, (c) Lasso regression cross validation error, (d) Prediction error for lasso regression, (e) Linear regression prediction plot, (e) Linear regression prediction plot, (f) Lasso regression prediction plot, (g) Ridge regression prediction plot and (h) MLP regressor prediction plot.

#### 5.2.3 Principal Component Analysis and Clustering Result

In Figures 13e and 10f, the first cluster has 40 countries, the second cluster 13 countries while the last cluster has only one country which is Botswana. We only show the two clusters dendrograms with many countries. In Figure 13c, average life expectancy has the highest positive correlation of 0.46 in PC 1 while first wave deterministic *R*_*o*_ has highest positive correlation in PC 2, value equal to 0.47.

**Figure 13.**
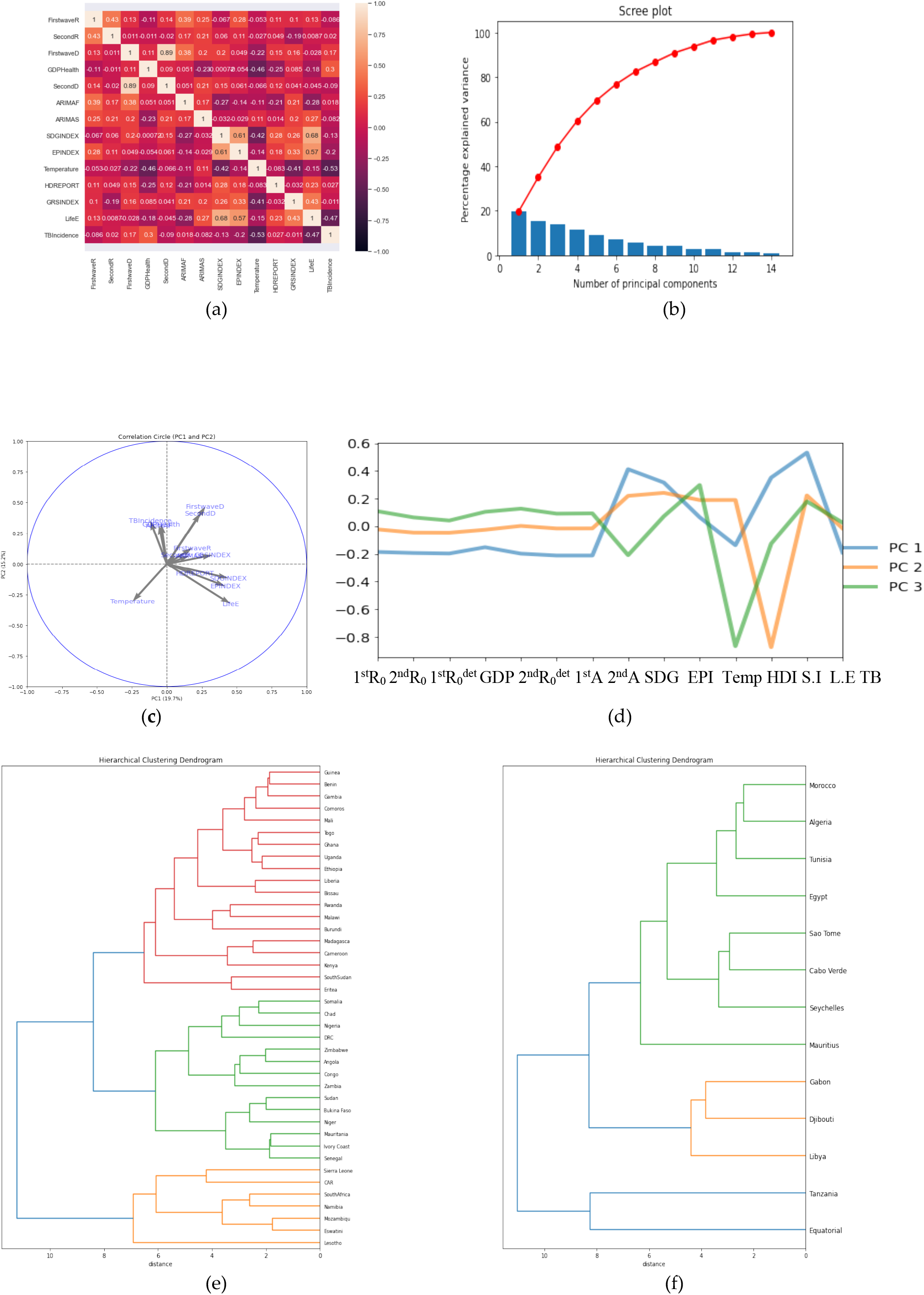
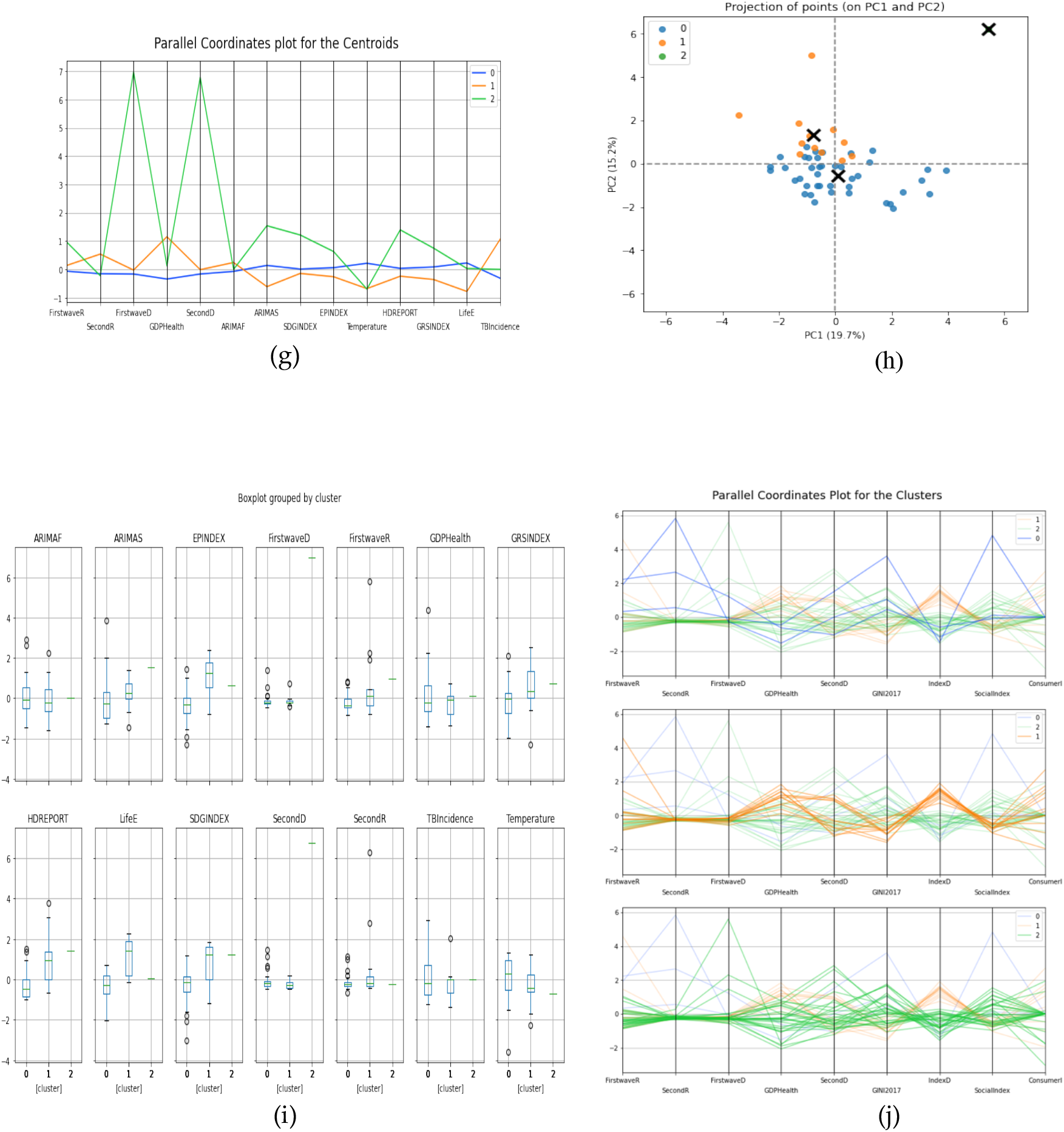
(a) Heatmap of the parameter’s correlations. (b) Scree plot. (c) & (d) PCs visualization. (e) & (f) Hierarchy clustering dendrograms. (g) Parallel coordinates plot for the centroids. (h) Projection points for PC1 and PC2. (i) Boxplot of the clusters. (j) Parallel coordinates plot for the clusters.

### 5.3 OECD Countries

#### 5.3.1 Multivariate Analysis for Socio-economic Variables and Epidemiologic Variables

Figure 14 corresponds to the ordinary multivariate least square method with R-squared = 0.90. Figure 15a shows Austria and Belgium as outliers not fitting the data.

**Figure 14.**
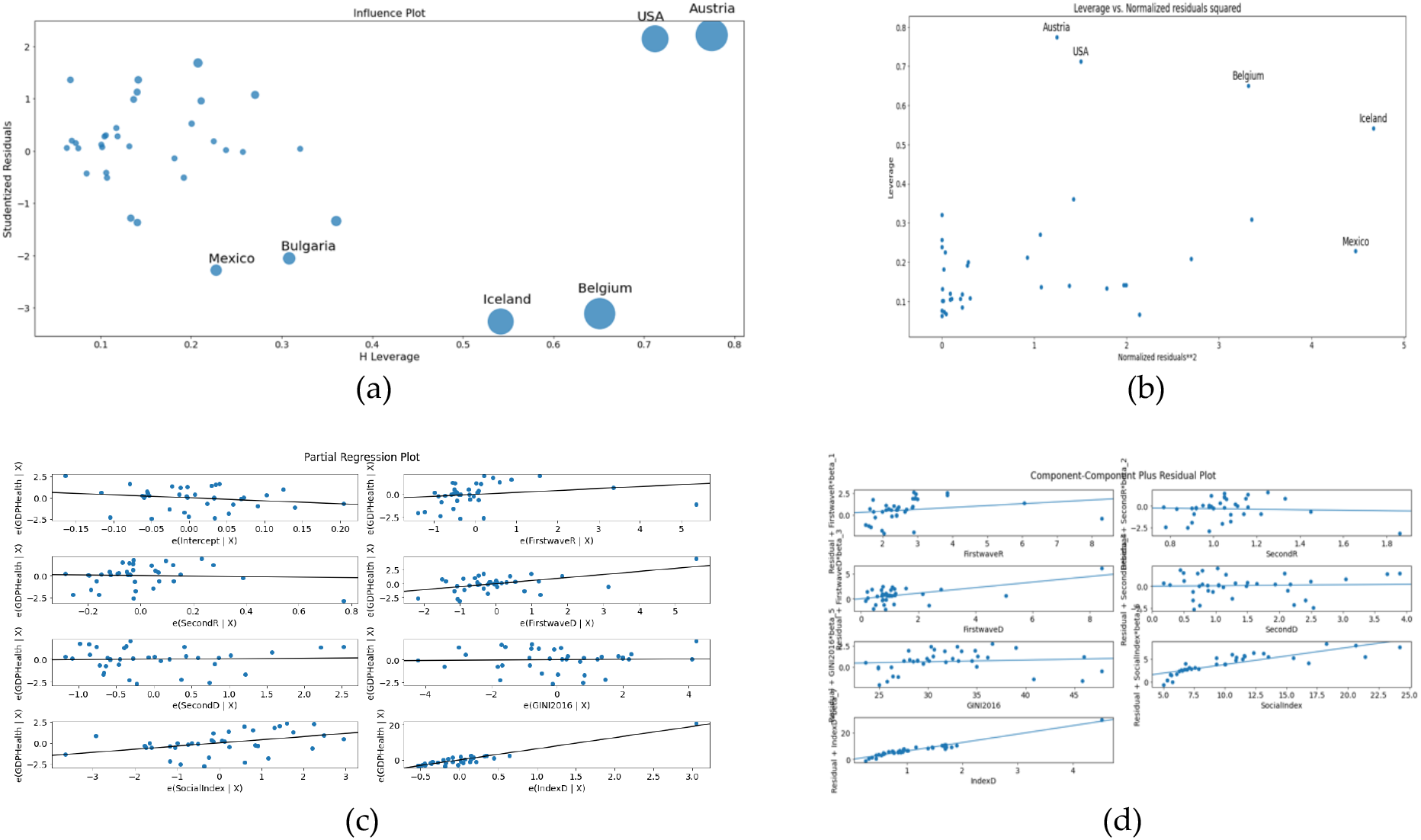
(a) Influence plot. (b) Leverage vs Normalized residuals squared plot. (c) Partial regression plot. (d) Component-Component plus residual plot.

**Figure 15.**
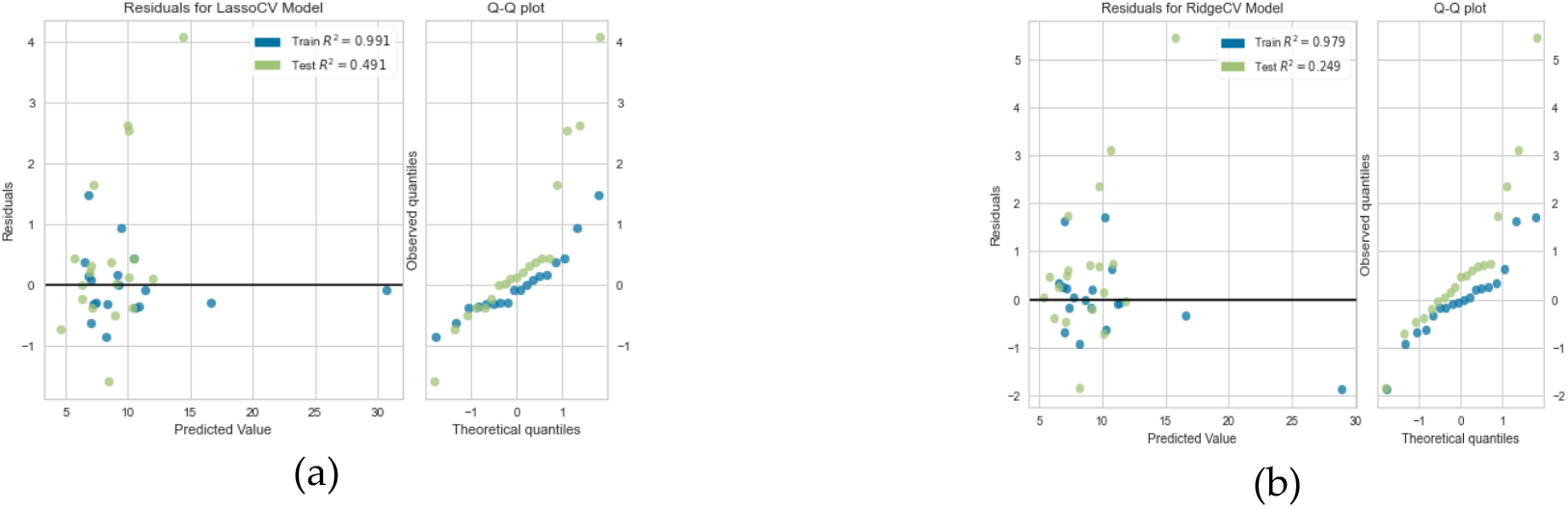
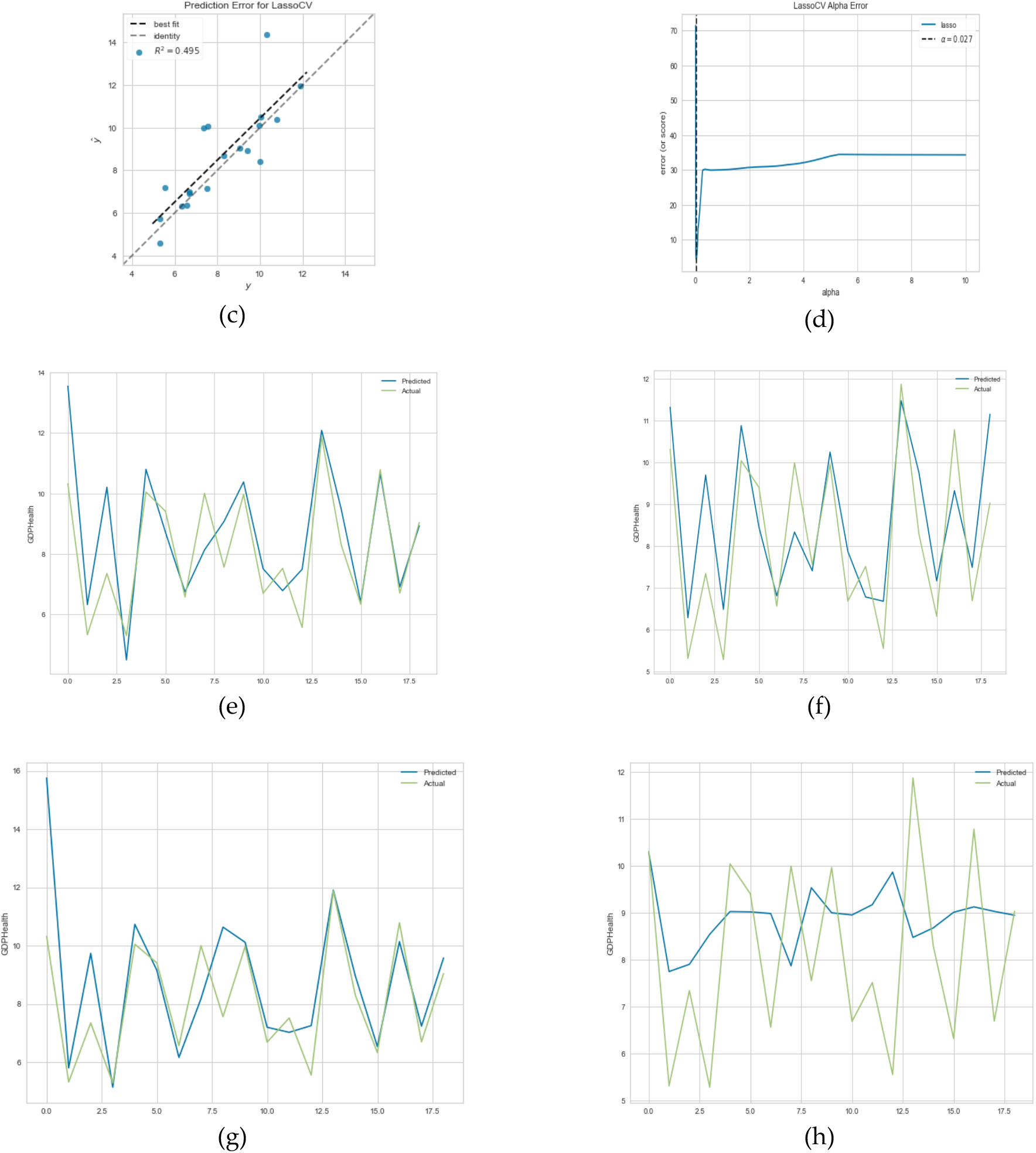
(a) Residual plot for lasso regression. (b) Residual plot for ridge regression. (c) Prediction error for lasso regression. (d) Lasso regression cross validation error. (e) Linear regression prediction plot. (e) Linear regression prediction plot. (f) Lasso regression prediction plot. (g) Ridge regression prediction plot. (h) MLP regressor prediction plot.

#### 5.3.2 Prediction of Percentage GDP Health Expenditure

In this section we used cross validation method to choose the best parameter α for the modelling as shown in Figure 15d. For ridge regression, α = 0.005 with mean square error of 1.905 and for Lasso regression, α = 0.027 with mean square error = 1.657. For Figure 15e, the training score = 0.993 and the test score = 0.535, for Figure 15f training score = 0.898 and the test score = 0.629, for Figure 15g training score = 0.983 and the test score = 0.259 and for Figure 15h training score = -0.072 and test score = -0.196. It is evident from these results that lasso regression best predicts percentage of GDP devoted to health expenditure with the highest test score and predicted values are very close.

#### 5.3.3 Principal Component Analysis and Clustering Result

In Figures 16e and 16f, the first cluster has 20 countries, the second has 5 countries which are USA and Bulgaria on same hierarchy, Mexico and Costa Rica on same hierarchy and Chile standing alone. The third cluster has 12 countries. We only show the two highest clusters dendrograms. In Figure 16c, Gini index and social fracture index have the highest positive correlation of 0.45 and 0.46 respectively in PC 1 while Gini index and percentage of GDP devoted to health expenditure has highest positive correlation in PC 2, whose values equal to 0.41 and 0.65 respectively.

**Figure 16.**
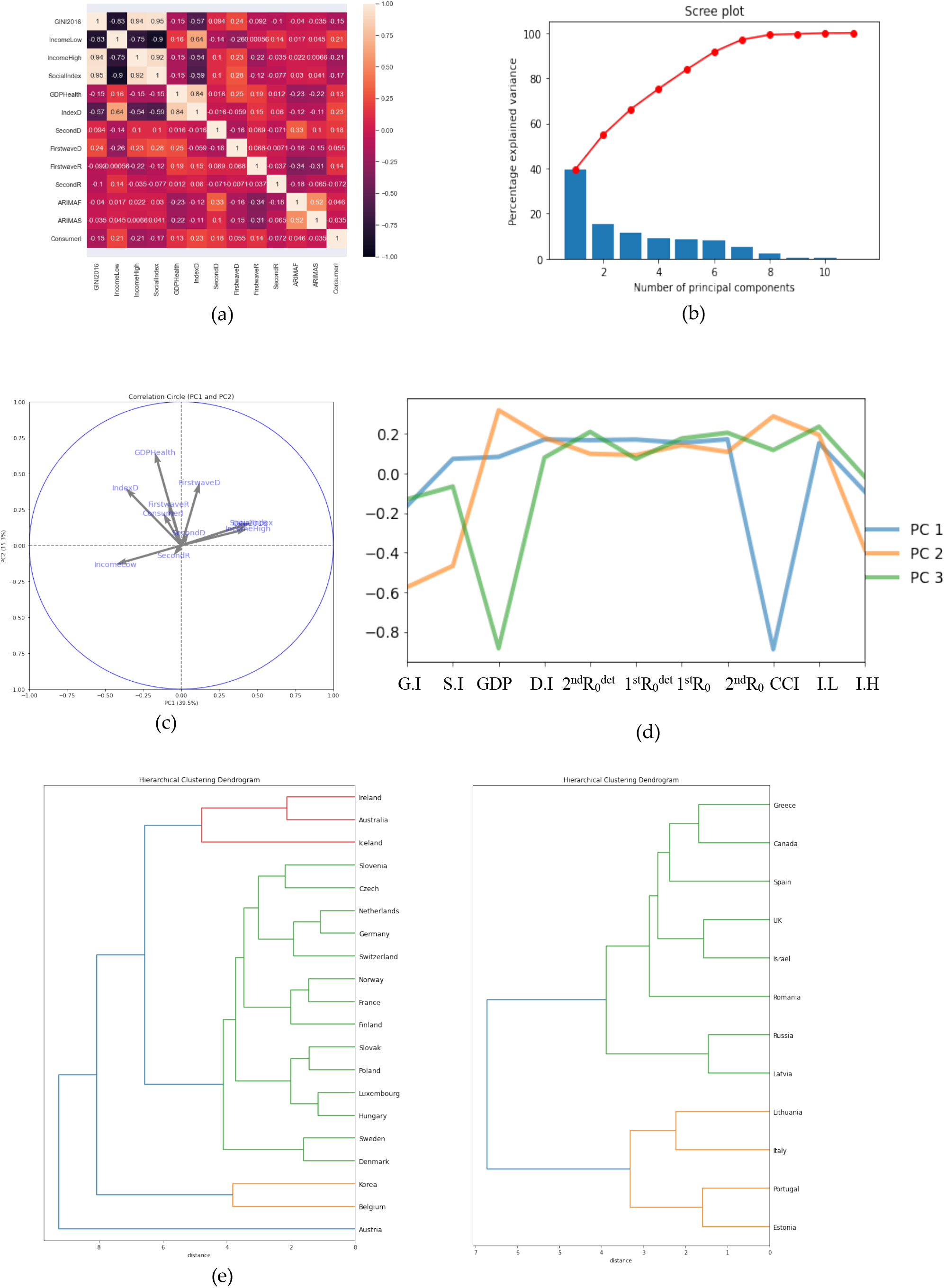
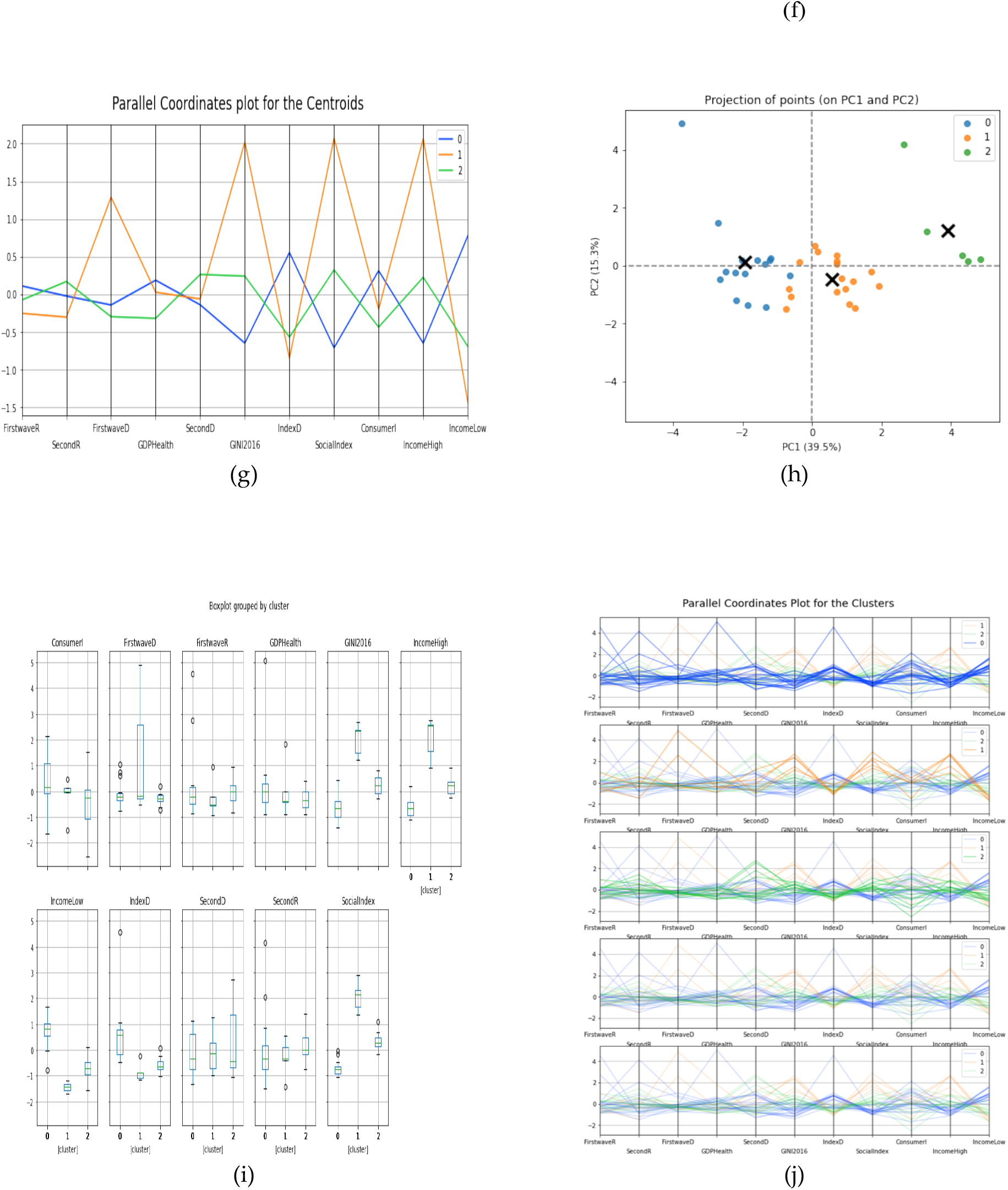
(a) Heatmap of the parameter’s correlations. (b) Scree plot. (c) & (d) PCs visualization. (e) & (f) Hierarchy clustering dendrogram. (g) Parallel coordinates plot for the centroids. (h) Projection points for PC1 and PC2. (i) Boxplot of the clusters. (j) Parallel coordinates plot for the clusters.

## Discussion

We have been able to develop new approaches to the socio-economic determinants for the modelling of the Covid-19 pandemic during the exponential phase. Some of these determinants have shown high correlation with parameters from epidemiology and can be seen in the heatmap diagrams in Figures 2, 10a, 13a, and 16a, explaining the role of each variable thanks to these correlations.

For developed and developing countries, lasso regression reduced the correlation between social fracture index and ten percent highest income to zero while for OECD countries Gini index and social fracture index correlation was reduced to zero. Some of our variables were not used in the optimization method-OLS due to multi-colinearity we observed at the summary of the results. For the two sets of countries, consumer confidence index, opposite initial autocorrelation averaged on six days for first and second wave, ten percent lowest income and ten percent highest income were not used in the modelling. The r-squared for OLS results for developed & developing and OECD countries are 0.76 and 0.90 respectively which shows a high significance rate (see figures 10e to 10f, 14).

The principal component analysis shows high correlation for the numbers of new cases we used in this research. Social fracture index has high correlation in PC1 for both cases while in PC2, percentage of GDP devoted to health expenditure was dominant for OECD countries and maximum *R*_+_ for the first wave was dominant for both developed and developing countries (see Figures 10c and 16c).

What we could deduce from all these observations is that the socio-economic determinants are a key to the modelling of infectious disease as these parameters give high signals on the trend during the spread of the pandemic for various countries.

## 7 Conclusion

The systematic study of the correlations between socio-economic variables (Gini and Theil indices, percentage of GDP devoted to heath expenditure, etc.) and epidemiological variables (reproduction rate, opposite of the slope of autocorrelation to origin, etc.) shows a disparity between developed and developing countries, as well as between epidemic waves. Developed countries with high indices of social divide, but high health expenditure, did not, for the first wave, react better to the COVID-19 epidemic than developing countries. On the other hand, the rapid implementation of isolation and vaccination measures enabled them to anticipate and reduce the effects of the second wave. In a subsequent work, we will study the evolution of this disparity between developed and developing countries during subsequent waves of SARS CoV-2.

## Data Availability

All data comes from public databases

## Authors Contributions

Conceptualization, J.D.; K.O. and M.R.; methodology, J.D.; K.O.and M.R.; software, K.O.; validation, J.D.; K.O. and M.R.; formal analysis, K.O.; investigation, J.D. and M.R.; resources, J.D.; data curation, K.O.; writing— original draft preparation, K.O.; writing—review and editing, J.D. and K.O; visualization, K.O.; supervision, J.D. and M.R; project administration, J.D. and M.R. All authors have read and agreed to the final version of the manuscript.

## Acknowledgments

The authors wish to acknowledge the Petroleum Technology Development Fund (PTDF) Nigeria doctoral fellowship in collaboration with Campus France Africa Unit.

## Appendix

**Table 3.** Developed and Developing Countries Data

| Country Name | 10% Lowest Income | 10% Highest Income | S.I Index | D.I | CCI | Gini Index | Country Name | Immigration Rate |
| --- | --- | --- | --- | --- | --- | --- | --- | --- |
| ALBANIA | 3.1 | 24.8 | 8.00 | 0.66 | - | 33.2 | AUSTRIA | 7.4 |
| AUSTRIA | 3.0 | 23.0 | 7.67 | 1.35 | 99.15 | 29.7 | BELARUS | 1.9 |
| BELGIUM | 3.3 | 21.9 | 8.11 | 1.27 | 100.75 | 27.4 | BELGIUM | 4.2 |
| BHUTAN | 2.7 | 27.9 | 10.33 | 0.30 | - | 37.4 | BRAZIL | 0.1 |
| BULGARIA | 1.9 | 31.9 | 16.79 | 0.44 | - | 40.4 | BULGARIA | -0.3 |
| CANADA | 2.7 | 25.3 | 9.37 | 1.15 | - | 33.3 | CANADA | 6.6 |
| CHILE | 2.3 | 36.3 | 15.78 | 0.58 | 96.55 | 44.4 | CHINA | -0.2 |
| CROATIA | 2.7 | 22.8 | 8.44 | 0.81 | - | 30.4 | CYPRUS | 4.2 |
| CYPRUS | 3.4 | 25.5 | 7.50 | 0.90 | - | 31.4 | CZECH | 2.1 |
| CZECH | 4.2 | 21.5 | 5.12 | 1.49 | 98.97 | 24.9 | DENMARK | 2.6 |
| DENMARK | 3.7 | 24.0 | 6.49 | 1.55 | 100.50 | 28.7 | ESTONIA | 0.3 |
| DJIBOUTI | 1.9 | 32.3 | 17.00 | 0.14 | - | 41.6 | FINLAND | 2.5 |
| EGYPT | 3.8 | 26.9 | 7.08 | 0.70 | - | 31.5 | FRANCE | 0.6 |
| ESTONIA | 3.0 | 22.5 | 7.50 | 0.89 | 97.59 | 30.4 | GERMANY | 6.6 |
| FINLAND | 3.8 | 22.6 | 5.95 | 1.52 | 100.27 | 27.4 | GREECE | -1.5 |
| FRANCE | 3.2 | 25.8 | 8.06 | 1.40 | 98.12 | 31.6 | HUNGARY | 1.3 |
| GABON | 2.2 | 27.7 | 12.59 | 0.22 | - | 38.0 | IRELAND | 4.9 |
| GREECE | 2.4 | 25.9 | 10.79 | 0.72 | 98.11 | 34.4 | ISRAEL | 1.2 |
| HUNGARY | 3.0 | 23.9 | 7.97 | 0.84 | 98.88 | 30.6 | ITALY | 2.5 |
| IRAN | 2.3 | 31.3 | 13.61 | 0.64 | - | 40.8 | KOSOVO | - |
| ITALY | 1.9 | 26.7 | 14.05 | 0.62 | 100.25 | 35.9 | LATVIA | -7.6 |
| KAZAKHSTAN | 4.3 | 23.0 | 5.35 | 0.55 | - | 27.5 | LITHUANIA | -11.6 |
| KOSOVO | 3.8 | 24.6 | 6.47 | - | - | 29.0 | LUXEMBOURG | 16.3 |
| LATVIA | 2.3 | 26.9 | 11.70 | 0.53 | 95.05 | 35.6 | MALTA | 2.1 |
| LESOTHO | 1.7 | 32.9 | 19.35 | 0.48 |  | 44.9 | MEXICO | -0.5 |
| LITHUANIA | 2.1 | 28.4 | 13.52 | 0.49 | 100.98 | 37.3 | NETHERLANDS | 0.9 |
| LUXEMBOURG | 2.4 | 25.8 | 10.75 | 0.49 | 98.52 | 34.9 | MARCEDONIA | -0.5 |
| MALTA | 3.4 | 23.3 | 6.85 | 1.31 | - | 29.2 | NORWAY | 5.3 |
| MAURITIUS | 2.9 | 29.9 | 10.31 | 0.57 | - | 36.8 | POLAND | -0.8 |
| MYANMAR | 3.8 | 25.5 | 6.71 | 0.71 | - | 30.7 | PORTUGAL | -0.6 |
| NETHERLAND | 3.5 | 23.3 | 6.66 | 1.50 | 99.93 | 28.5 | ROMANIA | -3.8 |
| MARCEDONIA | 1.7 | 23.8 | 14.00 | 0.47 | - | 34.2 | RUSSIA | 1.7 |
| NORWAY | 3.3 | 21.6 | 6.55 | 1.53 | - | 27.0 | SERBIA | 0.5 |
| POLAND | 3.2 | 23.5 | 7.34 | 0.86 | 98.70 | 29.7 | SLOVAK | 0.3 |
| PORTUGAL | 2.7 | 26.7 | 9.89 | 0.95 | 97.41 | 33.8 | SLOVENIA | 1.0 |
| ROMANIA | 1.6 | 24.9 | 15.56 | 0.36 | - | 36.0 | SPAIN | 0.9 |
| SERBIA | 1.4 | 25.6 | 18.29 | 0.47 | - | 36.2 | SWEDEN | 4.0 |
| SAO TOME | 1.3 | 49.1 | 37.77 | 0.17 | - | 56.3 | SWITZERLAND | 6.1 |
| SLOVENIA | 4.1 | 20.4 | 4.98 | 1.67 | 96.34 | 24.2 | TURKEY | 3.5 |
| SPAIN | 2.1 | 25.4 | 12.09 | 0.74 | 96.61 | 34.7 | UKRAINE | 0.2 |
| SWEDEN | 3.0 | 22.3 | 7.43 | 1.47 | 101.89 | 28.8 | UK | 3.9 |
| SWITZERLAND | 3.1 | 25.5 | 8.23 | 1.44 | 97.47 | 32.7 | USA | 2.9 |
| TANZANIA | 2.9 | 33.1 | 11.41 | 0.32 | - | 40.5 | - | - |
| ZIMBABWE | 2.5 | 34.8 | 13.92 | 0.34 | - | 44.3 | - | - |

**Table 4.** OECD Countries Data

| S/N | Country Name | 10% Lowest Income | 10% Highest Income | S.F Index | D.I | CCI | Gini Index |
| --- | --- | --- | --- | --- | --- | --- | --- |
| 1 | Australia | 2.8 | 26.1 | 9.32 | 1.00 | 100.86 | 32.5 |
| 2 | Austria | 3.3 | 22.5 | 6.82 | 4.52 | 99.15 | 28.0 |
| 3 | Belgium | 3.6 | 20.6 | 5.72 | 1.80 | 100.75 | 25.8 |
| 4 | Bulgaria | 1.9 | 31.9 | 16.79 | 0.44 | - | 40.8 |
| 5 | Canada | 2.6 | 24.2 | 9.31 | 1.16 | - | 30.3 |
| 6 | Chile | 1.8 | 37.1 | 20.61 | 0.44 | 96.55 | 46.0 |
| 7 | Costa Rica | 1.5 | 36.3 | 24.20 | 0.31 | 98.94 | 47.8 |
| 8 | Czech | 4.0 | 22.2 | 5.55 | 1.38 | 98.97 | 24.9 |
| 9 | Denmark | 4.0 | 21.2 | 5.30 | 1.90 | 100.50 | 26.4 |
| 10 | Estonia | 2.3 | 26.3 | 11.43 | 0.59 | 97.59 | 30.5 |
| 11 | Finland | 4.0 | 21.2 | 5.30 | 1.71 | 100.27 | 26.9 |
| 12 | France | 3.5 | 24.2 | 6.91 | 1.63 | 98.12 | 30.1 |
| 13 | Germany | 3.5 | 23.5 | 6.71 | 1.70 | 98.97 | 28.9 |
| 14 | Greece | 2.2 | 25.4 | 11.55 | 0.67 | 98.11 | 30.6 |
| 15 | Hungary | 3.1 | 22.5 | 7.26 | 0.92 | 98.88 | 28.9 |
| 16 | Iceland | 4.1 | 20.6 | 5.02 | 1.69 | - | 25.0 |
| 17 | Ireland | 3.1 | 24.4 | 7.87 | 0.88 | 100.48 | 29.5 |
| 18 | Israel | 2.0 | 26.3 | 13.15 | 0.57 | 99.15 | 34.8 |
| 19 | Italy | 2.1 | 24.4 | 11.62 | 0.75 | 100.25 | 33.4 |
| 20 | Korea | 2.2 | 22.0 | 10.00 | 0.76 | 99.43 | 34.5 |
| 21 | Latvia | 2.4 | 26.1 | 10.88 | 0.57 | 95.05 | 35.1 |
| 22 | Lithuania | 2.1 | 28.4 | 13.52 | 0.49 | 100.98 | 36.1 |
| 23 | Luxembourg | 3.5 | 22.1 | 6.31 | 0.84 | 98.52 | 31.8 |
| 24 | Mexico | 1.7 | 36.4 | 21.41 | 0.25 | 99.42 | 45.8 |
| 25 | Netherlands | 3.3 | 22.7 | 6.88 | 1.45 | 99.93 | 28.5 |
| 26 | Norway | 3.4 | 20.6 | 6.06 | 1.66 | - | 26.2 |
| 27 | Poland | 3.1 | 23.4 | 7.55 | 0.84 | 98.70 | 28.1 |
| 28 | Portugal | 2.4 | 26.2 | 10.92 | 0.86 | 97.41 | 31.7 |
| 29 | Romania | 1.6 | 24.9 | 15.56 | 0.36 | - | 35.0 |
| 30 | Russia | 2.9 | 29.1 | 10.03 | 0.53 | 96.12 | 33.1 |
| 31 | Slovak | 3.3 | 21.6 | 6.55 | 1.02 | 97.71 | 23.6 |
| 32 | Slovenia | 3.6 | 20.4 | 5.67 | 1.46 | 96.34 | 24.9 |
| 33 | Spain | 2.0 | 24.7 | 12.35 | 0.73 | 96.61 | 33.0 |
| 34 | Sweden | 3.5 | 22.6 | 6.46 | 1.69 | 101.89 | 27.5 |
| 35 | Switzerland | 3.4 | 24.1 | 7.09 | 1.68 | 97.47 | 29.9 |
| 36 | UK | 2.7 | 28.6 | 10.59 | 0.94 | 98.71 | 36.6 |
| 37 | USA | 1.6 | 29.2 | 18.25 | 0.93 | 98.68 | 39.0 |

**Table 5.** AFRICA Countries Data

| S/N | Country Name | SDG Index | EPI | Temperature | HDI | S.I | L.E | TB Incidence |
| --- | --- | --- | --- | --- | --- | --- | --- | --- |
| 1 | Algeria | 65.8 | 57.18 | 18.79 | 21 | 72.22 | 78.0 | - |
| 2 | Angola | 49.3 | 37.44 | 21.55 | - | 58.33 | 61.0 | 3.51 |
| 3 | Benin | 51.5 | 38.17 | 27.55 | 3.2 | 43.52 | 61.0 | 0.55 |
| 4 | Botswana | 61.6 | 51.70 | 21.50 | 20 | 62.04 | 65.0 | - |
| 5 | Bukina Faso | 53.5 | 42.83 | 28.29 | 0.6 | 22.22 | 63.0 | 0.47 |
| 6 | Burundi | 50.3 | 27.43 | 20.80 | - | 20.37 | 67.0 | 1.07 |
| 7 | Cabo Verde | 64.1 | 56.94 | 23.30 | 19 | 45.37 | 73.0 | 0.46 |
| 8 | Cameroon | 51.6 | 40.81 | 24.60 | 1.4 | 35.19 | 62.0 | 1.79 |
| 9 | CAR | 36.7 | 36.42 | 24.90 | 2.9 | 18.52 | 54.0 | 5.40 |
| 10 | Chad | 38.7 | 45.34 | 26.55 | - | 22.22 | 58.0 | 1.42 |
| 11 | Comoros | 47.6 | 44.24 | 25.55 | 4.2 | - | 66.0 | 0.35 |
| 12 | DRC | 41.6 | 30.41 | 24.00 | 0.1 | - | 61.0 | 3.20 |
| 13 | Congo | 48.7 | 42.39 | 24.55 | - | 47.22 | 61.0 | 3.73 |
| 14 | Ivory Coast | 55.6 | 45.25 | 26.35 | 21 | 25.93 | 61.0 | 1.37 |
| 15 | Djibouti | 49.7 | 40.04 | 28.00 | 40 | 37.96 | 65.0 | - |
| 16 | Egypt | 63.8 | 61.21 | 22.10 | 3.6 | 54.63 | 74.0 | - |
| 17 | Equatorial | 43.4 | 60.40 | 24.55 | - | - | 66.0 | 1.81 |
| 18 | Eritea | 43.3 | 39.34 | 25.50 | 0.1 | 86.11 | 66.0 | 0.86 |
| 19 | Eswatini | 52.4 | - | 21.40 | 5.0 | 68.52 | 59.0 | 3.63 |
| 20 | Ethiopia | 53.2 | 44.78 | 22.20 | 8.4 | 50.93 | 68.0 | 1.40 |
| 21 | Gabon | 59.4 | 45.05 | 25.05 | 20 | 50.93 | 69.0 | 5.21 |
| 22 | Gambia | 51.9 | 42.42 | 27.50 | 4.3 | 44.44 | 66.0 | 1.58 |
| 23 | Ghana | 61.2 | 49.66 | 27.20 | 2.5 | 50.93 | 68.0 | 1.44 |
| 24 | Guinea | 49.4 | 46.62 | 25.70 | 3.1 | 52.78 | 63.0 | 1.76 |
| 25 | Bissau | 45.5 | 44.67 | 26.75 | 1.3 | - | 63.0 | 3.61 |
| 26 | Kenya | 56.6 | 47.25 | 24.75 | 0.9 | 50.93 | 69.0 | 2.67 |
| 27 | Lesotho | 50.9 | 33.78 | 11.85 | 6.8 | 65.74 | 53.0 | 6.54 |
| 28 | Liberia | 48.0 | 41.62 | 25.30 | - | 40.74 | 65.0 | 3.08 |
| 29 | Libya | - | 49.79 | 21.80 | 34 | 76.85 | 77.0 | - |
| 30 | Madagasca | 45.6 | 33.73 | 22.65 | 4.9 | 37.04 | 67.0 | 2.33 |
| 31 | Malawi | 52.3 | 49.21 | 21.90 | 1.9 | 48.15 | 63.0 | 1.46 |
| 32 | Mali | 51.7 | 43.71 | 28.25 | 2.5 | 48.15 | 62.0 | 0.52 |
| 33 | Mauritania | 51.3 | 39.24 | 27.65 | 14 | 29.63 | 65.0 | 0.89 |
| 34 | Mauritius | 66.2 | 56.63 | 22.40 | 3.0 | 93.52 | 76.0 | 0.12 |
| 35 | Morocco | 64.4 | 63.47 | 18.10 | 19 | 76.85 | 73.0 | - |
| 36 | Mozambiqu | 51.4 | 46.37 | 23.80 | 5.1 | 55.56 | 56.0 | 3.61 |
| 37 | Namibia | 57.1 | 58.46 | 20.95 | 1.4 | 42.59 | 65.0 | 4.86 |
| 38 | Niger | 50.3 | 35.74 | 27.15 | 7.3 | 34.26 | 59.0 | 0.84 |
| 39 | Nigeria | 47.1 | 54.76 | 26.80 | 0.1 | 45.37 | 60.0 | 2.19 |
| 40 | Rwanda | 57.9 | 43.68 | 18.85 | 1.8 | 71.30 | 65.0 | 0.57 |
| 41 | Sao Tome | 61.8 | 54.01 | 23.75 | - | - | 66.0 | 1.14 |
| 42 | Senegal | 57.0 | 49.52 | 27.85 | 12 | 13.89 | 63.0 | 1.17 |
| 43 | Seychelles | - | 66.02 | 27.15 | - | 65.74 | 76.0 | 0.16 |
| 44 | Sierra Leone | 49.7 | 42.54 | 23.82 | 6.6 | 48.15 | 60.0 | 2.95 |
| 45 | Somalia | 40.1 | - | 27.05 | - | 47.22 | 54.0 | 2.58 |
| 46 | SouthAfrica | 60.4 | 44.73 | 18.75 | 1.0 | 48.15 | 65.0 | 6.15 |
| 47 | SouthSudan | 29.2 | - | - | - | 72.22 | - | 2.27 |
| 48 | Sudan | 47.4 | 51.49 | 26.90 | 5.4 | 24.07 | 67.0 | 0.67 |
| 49 | Tanzania | 55.9 | 50.83 | 22.35 | 16 | 8.33 | 64.0 | 2.37 |
| 50 | Togo | 52.7 | 41.78 | 27.15 | 1.1 | 67.59 | 67.0 | 0.37 |
| 51 | Tunisia | 66.1 | 62.35 | 16.30 | 5.4 | 62.04 | 76.0 | - |
| 52 | Uganda | 54.9 | 44.28 | 22.80 | 0.5 | 53.70 | 68.0 | 2.00 |
| 53 | Zambia | 53.0 | 50.97 | 21.40 | 20 | 36.11 | 54.0 | 3.33 |
| 54 | Zimbabwe | 54.8 | 43.41 | 21.00 | 16 | 53.70 | 62.0 | 1.99 |

